# The influence of COVID-19 risk perception and vaccination status on the number of social contacts across Europe: insights from the CoMix study

**DOI:** 10.1101/2022.11.25.22282676

**Authors:** James Wambua, Neilshan Loedy, Christopher I Jarvis, Kerry LM Wong, Christel Faes, Rok Grah, Bastian Prasse, Frank Sandmann, Rene Niehus, Helen Johnson, W.John Edmunds, Philippe Beutels, Niel Hens, Pietro Coletti

## Abstract

**Background:** The SARS-CoV-2 transmission dynamics have been greatly modulated by human contact behaviour. To curb the spread of the virus, global efforts focused on implementing both Non-Pharmaceutical Interventions (NPIs) and pharmaceutical interventions such as vaccination. This study was conducted to explore the influence of COVID-19 vaccination status and risk perceptions related to SARS-CoV-2 on the number of social contacts of individuals in 16 European countries. This is important since insights derived from the study could be utilized in guiding the formulation of risk communication strategies.

**Methods:** We used data from longitudinal surveys conducted in the 16 European countries to measure social contact behaviour in the course of the pandemic. The data consisted of representative panels of participants in terms of gender, age and region of residence in each country. The surveys were conducted in several rounds between December 2020 and September 2021. We employed a multilevel generalized linear mixed effects model to explore the influence of risk perceptions and COVID-19 vaccination status on the number of social contacts of individuals.

**Results:** The results indicated that perceived severity played a significant role in social contact behaviour during the pandemic after controlling for other variables. More specifically, participants who perceived COVID-19 to be a serious illness made fewer contacts compared to those who had low or neutral perceptions of the COVID-19 severity. Additionally, vaccinated individuals reported significantly higher number of contacts than the non-vaccinated. Furthermore, individual-level factors played a more substantial role in influencing contact behaviour than country-level factors.

**Conclusion:** Our multi-country study yields significant insights on the importance of risk perceptions and vaccination in behavioral changes during a pandemic emergency. The apparent increase in social contact behaviour following vaccination would require urgent intervention in the event of emergence of an immune escaping variant. Hence, insights derived from this study could be taken into account when designing, implementing and communicating COVID-19 interventions.

## Background

The COVID-19 pandemic has led to unprecedented wide-ranging effects across the globe on social life and mental health, and has adversely affected the global economy [1, 2, 3]. Since its emergence, it has led to massive loss of lives worldwide with an estimated 6.5 million confirmed COVID-19 deaths as of November, 21st 2022 [4], and an approximated 14.9 million excess deaths associated with the pandemic in 2020 and 2021 [5]. Furthermore, there is mounting evidence that some of the people who suffered from the COVID-19 disease experience prolonged adverse health effects with continued multisystemic symptoms weeks and months post infection with a substantial impact on health and wellbeing [6, 7]. From March 11, 2020, the time COVID-19 was declared a global pandemic by the World Health Organization [8], different countries have experienced different waves of the pandemic. This in turn has led to implementation of a range of interventions to alleviate pressure on the healthcare systems as well as to control the pandemic [9, 10]. Before the introduction of the COVID-19 vaccines, governments relied on adopting various Non-Pharmaceutical Interventions (NPIs) to curtail the transmission dynamics of SARS-CoV-2 [11]. Since the introduction of the COVID-19 vaccines, different governments have adopted different vaccine roll out strategies. As vaccine uptake is increasing globally [12], governments are monitoring and adjusting NPIs depending on the epidemic situation in their respective countries [10]. Due to the continuing uncertainty on the future trajectory of the pandemic, as has been demonstrated by the newly emerging COVID-19 variants of concern such Omicron and its sub-variants [13, 14], global efforts on both vaccination and proportionate implementation of relevant NPIs are essential to reduce cases without too much negative social and economic impacts [15].

Since NPIs mainly encompass social distancing measures, adherence to these measures might be influenced by attitudinal and demographic determinants [16]. During the past pandemics, the control of fast spreading infectious diseases has relied, partially on populations’ risk perceptions, both at individual and societal level [17]. Thus, investigating the key attitudinal determinants influencing behavioral responses is pivotal to continue guiding the implementation of appropriate strategies. Risk perception is a key component of the Health Belief Model (HBM) [18] and the theory of Protection Motivation and Self-efficacy (PMS) [19]. The HBM framework emphasizes that individual’s likelihood to adopt health preventive behaviours are mainly based on their risk perceptions [18]. Whilst the PMS theory postulates that the implementation of the recommended health protective behaviours is based on individual’s risk perceptions and self-efficacy to adopt them [19]. Several empirical studies have explored the relationship between risk perceptions and the adoption of health protective measures during the current COVID-19 pandemic [20, 21, 22, 23, 24, 25, 26] and previous pandemics [27, 28] and found that risk perceptions play a significant role in the adoption of health protective measures. The most utilized health protective behaviours in these studies include physical distancing, frequency of hand washing, wearing of face masks, and avoiding public places. Risk perception has yielded significant relationships with the number of social contacts from two recent studies, one in Belgium [26] and the other in UK [29] during the COVID-19 pandemic.

In addition to the NPIs which are mainly focused on reducing close person-to-person contacts, following the initiation and continued uptake of COVID-19 vaccination, it is imperative to explore whether vaccination alters social contact behaviour given the inherent uncertainties in vaccine waning as well as protection against emerging variants [30]. However, the literature on the relationship between COVID-19 vaccination status and perceptions and social contact behaviour during a pandemic is very limited. Thus, given the crucial role of contact behaviour in the dynamics of SARS-CoV-2 virus [31, 32], it is important to analyse how specific COVID-19 perceptions and vaccination status relate to the number of social contacts in a wider geographical context for the ongoing management of the pandemic. Utilizing data collected under different phases of the pandemic and also under different intervention measures from multiple countries is crucial to correctly disentangle possible transient and/or country specific effects.

Here, we present analyses of the influence of COVID-19 risk perceptions and vaccination status on the number of social contacts of individuals using longitudinal social contact data collected during the COVID-19 pandemic as part of the CoMix study [33]. This is pivotal to continue enhancing the understanding between risk perceptions and social contacts as the world continue pushing towards a post-acute phase of the pandemic.

## Methods

### Ethics statement

Approvals or waivers for the CoMix study questionnaires and protocols were obtained through the local ethical committees for each individual country in the study. Participants aged 18 years and older opted to voluntarily participate in the study. All the analyses were performed on pseudo-anonymised data. Informed consent was obtained from all subjects involved in the study. The country-level ethical details or waivers are described in more details in Verelst et al [33].

#### 0.1 Survey Methodology

The CoMix study is an online multi-country longitudinal social contact survey conducted during the COVID-19 pandemic [33]. The survey started in March 2020 in the UK [34] and April 2020 in the Netherlands [35] and Belgium [31]. Between December 2020 and April 2021, the survey was extended to eight additional European countries (Denmark, Austria, France, Poland, Italy [36], Portugal, Poland and Spain). Between February 2021 and October 2021, the survey was further extended to eight more European countries (Greece, Finland, Hungary, Estonia, Slovakia, Lithuania, Switzerland and Croatia). In this paper, we used data from the latter 16 countries, pertaining to surveys taken between December 2020 and September 2021. For more details on the country-specific timelines of the data collection for each survey round, see Supplementary Table 1, Additional file 1. In each of these countries, representative panels of participants aged 18 years or above were invited to complete the CoMix survey. The panels of participants in each country were nationally-representative based on gender, age, and region of residence. Although the survey collected information in adults, data for children (i.e. below 18 years of age) was collected via a proportion of the adult respondents reporting for only one chosen child in their household. The recruitment and data collection were conducted by a contracted market research company. In each country, the data was collected from the same individuals in successive survey waves. In each survey wave, individuals who agreed to participate were asked to report retrospectively the number of social contacts made between 5am on the day preceding the survey day and 5am of the survey day. A social contact was defined as any conversation the participant had in person which involved at least an exchange of a few words, or involved a skin-to-skin contact. In addition, participants provided demographic information such as age, gender, high-risk status, household size, socio-economic status, data related to risk perceptions, information on whether the participant had received vaccination against the COVID-19.

The risk perceptions consisted of three statements that the participants were asked to respond to. The first statement was *“I am likely to catch coronavirus”*, the second was *“I am worried that I might spread coronavirus to someone who is vulnerable”*, and the third was *“Coronavirus would be a serious illness for me”*. The responses to the statements were coded on 5-point Likert scales: strongly agree, tend to agree, neither agree nor disagree, tend to disagree and strongly disagree for reliability analysis. We used Cronbach’s alpha [37] as a measure for the internal consistency and the results yielded three separate constructs (see Supplementary Section 0.1 on Reliability analysis, Additional file 1). We refer to these constructs as, *perceived susceptibility, perceived risk to the vulnerable* and *perceived severity* for the first, second and third items, respectively. Statistical analysis was based on participants aged 18 years or above and considering three response levels: low perception (‘tend to disagree’ and ‘strongly disagree’), neutral (‘neither agree nor disagree’ and ‘don’t know’) and high perception (‘strongly agree’ and ‘tend to agree’).

#### 0.2 Data on stringency index

Due to the wide variations in the intervention measures initiated over time, it was important to standardize the stringency of local policy choices. This led to the launch of Oxford COVID-19 Government Response Tracker (OxCGRT), which provides a unified approach to follow the different government responses in different countries, and in some instances, sub-national restrictions over time [10]. This project utilizes a series of standardized indicators which are summed up to create composite indexes to represent the implementation levels of different intervention measures. The stringency index is computed using the following containment and closure policies (closure of schools and universities, closure of workplaces, limitations on gathering sizes, cancellation of public events, closure of public transport, restrictions to stay at home, and restrictions on domestic and international travels/movements). The stringency index takes values ranging from 0 (least stringent) to 100 (most stringent), and allows for cross-national comparisons of the implemented measures and policies. The OxCGRT is updated regularly as the government response measures change. This data is mainly obtained from publicly available sources such as government’s briefings, mass articles, press releases among others. More information can be obtained from Hale et al [10].

## Statistical Analyses

We employed a multilevel generalized linear mixed effects model (GLMM) to explore the associations between the risk perceptions and vaccination status with the number of social contacts [38]. The participants observations from each survey round (level-1) were nested within participants (level-2), and participants were nested within countries (level-3). Random effects were used to correct for correlations in this nested nature of the study. We also performed exploratory modeling using GLMM where the country was considered as a fixed effect and the participants as random effects to explore cross-level effects of the country and individual perception variables [39]. In both modeling approaches, the number of social contacts is modelled using (1) a negative binomial distribution, accounting for possible overdispersion in the counts, and (2) zero-inflation component to deal with excess zeroes in the number of social contacts. The intra-class correlation, which is a quantity measure of the between and within group variation [40], was used to gain insights on heterogeneity in number of social contacts between and within the countries. Throughout our analyses, the number of contacts were truncated at 100. The models included the individual risk perception variables and vaccination status (vaccinated versus not vaccinated) and adjusted for the participant’s household size, gender, age, day of the week (week day versus weekend), self reported high risk status, history of COVID-19 infection, employment status, and stringency index as potential confounders. The stringency index was considered in four levels: low (0-40), moderate (41-55), high (56-70) and very high (71-100). The vaccination status was primarily defined based on the first dose of any of the COVID-19 vaccines since some surveys were conducted in the initial phases of the vaccination where few individuals were fully vaccinated. History of infection was defined based on whether participants had previously tested for the virus. The models were fitted using maximum likelihood estimation. We used R version 4.1.1 and the glmmTMB package (version 1.0.2.1) [41] for all statistical analyses.

Model building was performed for each individual perception variable. This was informed by preliminary exploratory analyses of a model including all the perception variables and models for each individual perception variable. Hence in total, we had 3 models for the multilevel generalized linear mixed effects corresponding to each individual perception variable (i.e. *perceived severity, perceived susceptibility, perceived risk to vulnerable*) and 3 models for the generalized linear mixed effects including the country as a fixed effect. The significance of the model variables was assessed through Type III Wald tests and a significance level of 5% was considered.

## Results

### Descriptive results

The final sample size included in the analysis consisted of 29,292 participants aged 18 years or above from the 16 countries for a total of 111,103 completed surveys. Among these, 50.1% were completed by males, 49.7% by females, and 0.2% by participants who did not indicate their gender. Age was categorized in six age groups, with 15.5% participants in age group (18 - 29), 17.2% in age group (30 - 39), 19.2% in age group (40 - 49), 18.7% in age group (50 - 59), 19.5% in age group (60 - 69), and 9.7% in age group (70 - 120). The mean number of surveys each participant completed was 3.9 in the aggregated dataset, with a range of 3.2 - 4.4 when looking at individual countries data, with a maximum of 7 waves. More information on sample characteristics for each country is contained in Supplementary Table 2, Additional file 1.

We observed noticeable differences in the levels of risk perceptions in the different countries with slight variations over time (Figure 1). The comparison of the three risk perceptions indicated that in general, the perceived risk to the vulnerable was the highest followed by perceived severity and perceived susceptibility, respectively. The percentages of the vaccinated participants showed an increasing trend during the study period (Supplementary Figure 1, Additional file 1). The level of stringency of intervention measures showed apparent differences between countries and little variations within each individual country during the study period (Supplementary Figure 2, Additional file 1).

**Figure 1:**
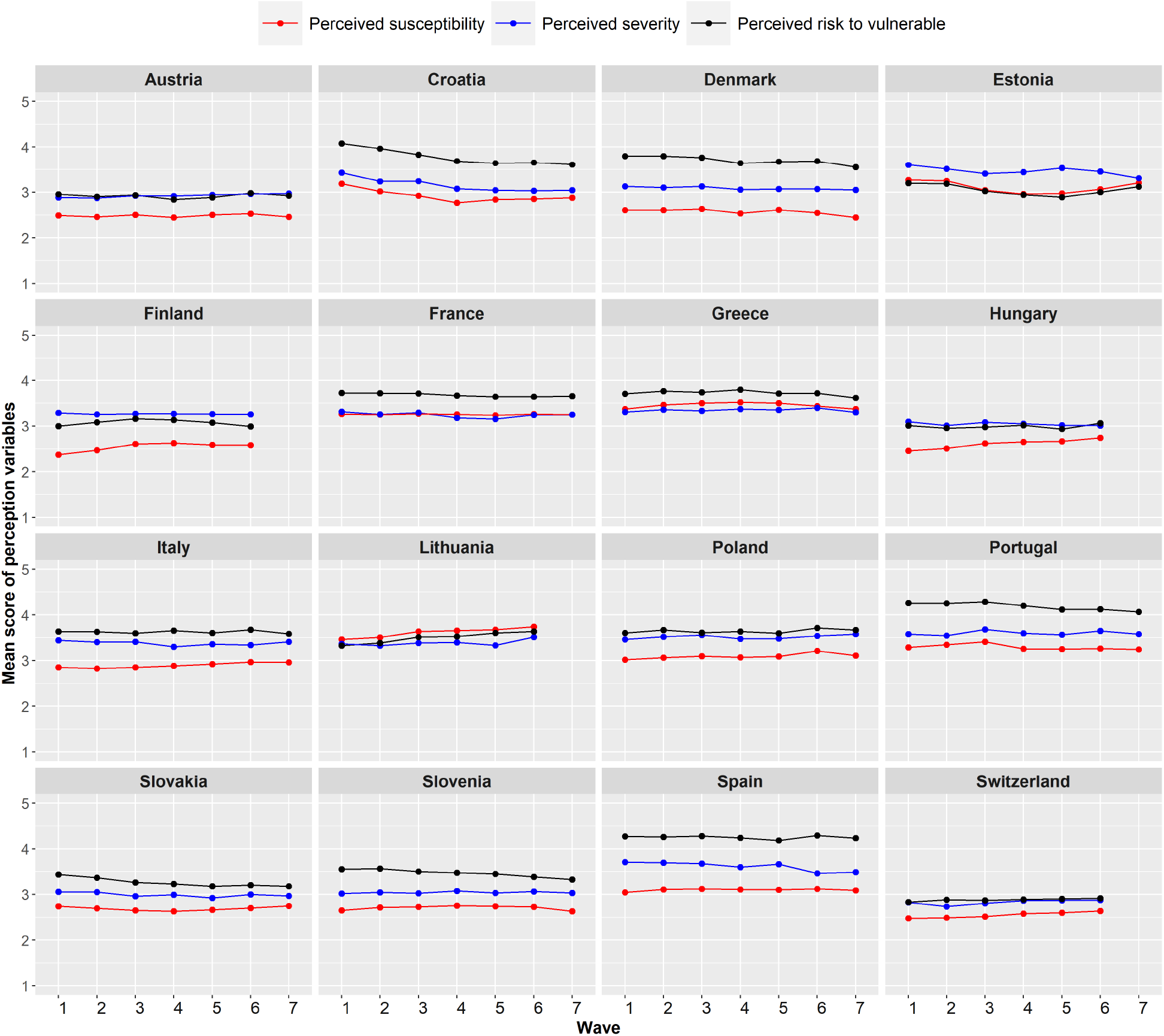
Mean score of perceived severity, perceived susceptibility and perceived risk to the vulnerable during the data collection period in the different countries. The Likert scale was assigned as follows;”Strongly agree” = 5,”Tend to agree” = 4,”Neither agree nor disagree” = 3,”Tend to disagree” = 2,”Strongly disagree” = 1.

### Multilevel GLMM model with country as a random effect

#### Summary results

Results from the multilevel generalized linear mixed effects model (GLMM) indicated that the overdispersion parameter ranged between 9.29 (95% CI 6.80 - 12.70) and 9.36 (95% CI 6.87 - 12.77) in the different models indicating substantial overdispersion in the number of social contacts. The nested random effect of participants within countries was statistically significant from the likelihood ratio test (p-value *<*0.001), further indicating considerable heterogeneity in social contact behaviour between individuals. The total variance in the number of social contacts from the individual participants (level-2) of the nested random effects was 0.627 (95% CI 0.594 - 0.659). Whilst the total variance from the countries (level-3) was 0.012 (95% CI 0.006 - 0.0267). The intra-class correlation was 1.9% in our models. This suggests substantial variability in social contact behaviour between individuals within the same country and low variability in social contact behaviour between countries.

#### Perceived severity

Results from the model for the perceived severity indicated that participants with low levels of perceived severity reported 1.25 (95% CI 1.13 - 1.37) times more contacts than participants who had high levels of perceived severity after controlling for the other factors (Supplementary Table 3, Additional file 1). Similarly, the model results showed that participants who had neutral perceptions on the severity of COVID-19 reported 1.10 (95% 1.00 - 1.21) times more social contacts than those who had high levels of perceived severity (Supplementary Table 3, Additional file 1). The predicted mean number of contacts for participants with high levels of perceived severity was 2.21 (95% CI 1.95 - 2.51), whilst for those with neutral or low perception on severity was 2.43 (95% CI 2.14 - 2.76) and 2.76 (95% CI 2.44 - 3.12), respectively. These mean predicted number of contacts in the period between December 2020 and September 2021 are relatively much lower compared to the average number of contacts reported in the prepandemic period in a multi-country social contact survey (POLYMOD) conducted in 8 European countries [42]. These countries included Belgium, Finland, Germany, Italy, Luxembourg, Netherlands, Poland and UK. The average number of contacts was 13.4 with country-level mean contacts ranging between 7.95 (standard deviation (SD) 6.26) and 19.77 (SD 12.27), with the lowest reported in Germany and the highest in Italy.

Furthermore, the results for the perceived severity model yielded significant interaction effects between perceived severity and vaccination status (p-value = 0.011) (Supplementary Table 4, Additional file 1). In both the vaccinated and not vaccinated groups, participants who had high levels of perceived severity reported fewer number of social contacts in comparison with participants who had either low or neutral levels of perceived severity (Figure 2). Overall, vaccinated individuals reported on average 1.31 (95% CI 1.23 - 1.39) times more contacts than the non vaccinated. The predicted number of contacts for the vaccinated was 2.89 (95% CI 2.56 - 3.27), whilst for the non-vaccinated was 2.21 (95% CI 1.95 - 2.51). We also considered the predicted number of contacts from the marginal effects of the interactions terms between perceived severity and stringency index. We observed that the differences in predicted number of contacts among the perceived severity levels were more pronounced for the low levels of stringency index. (Figure 3**a**). Moreover, considering the predicted number of contacts from the marginal effects of the interaction terms between perceived severity and age group, more variation in the predicted number of contacts was observed in higher age groups (Figure 3**b**).

**Figure 2:**
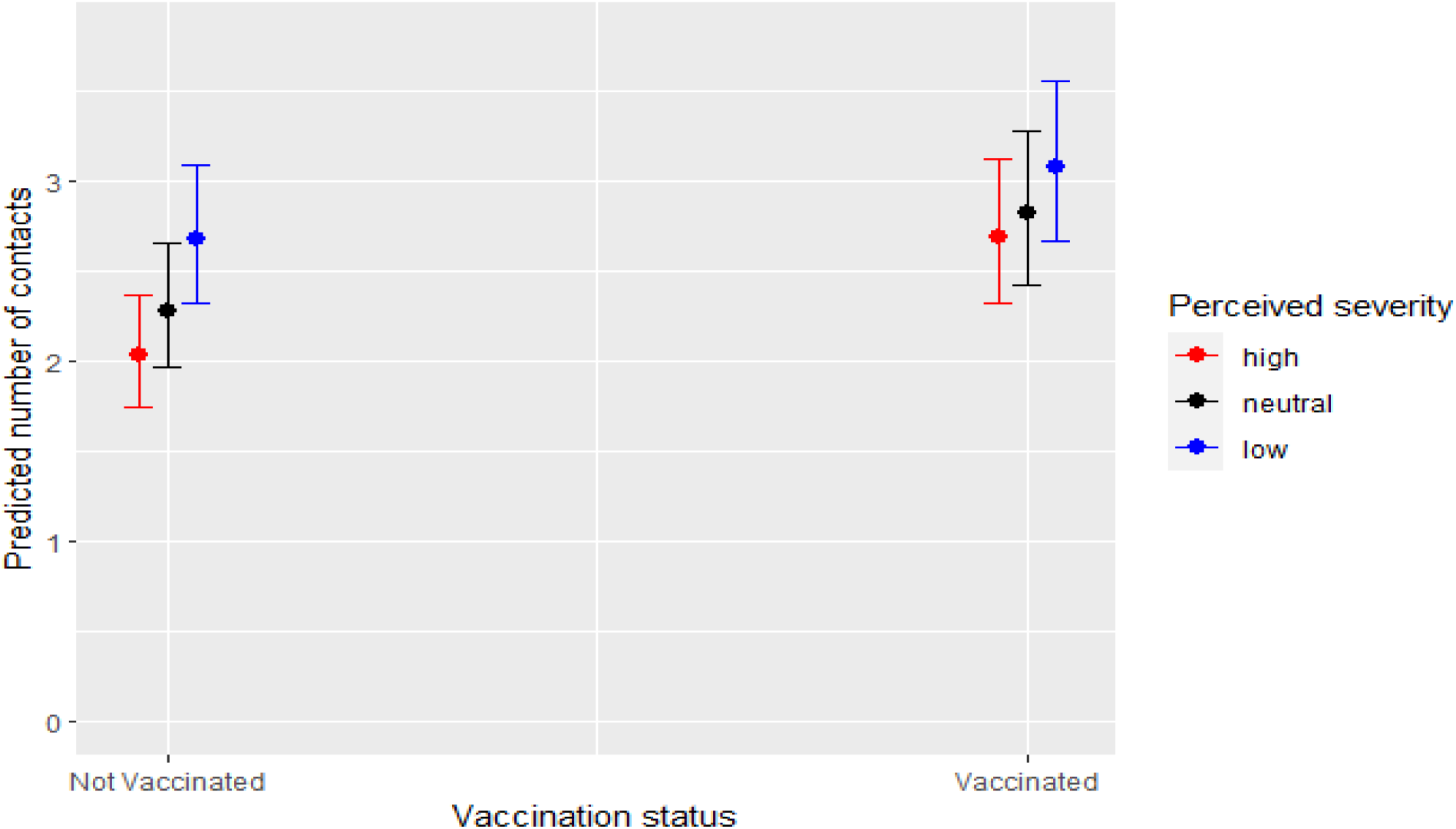
Predicted number of contacts from the perceived severity model with 95% confidence interval (CI) in the period between December 2020 and September 2021.

**Figure 3:**
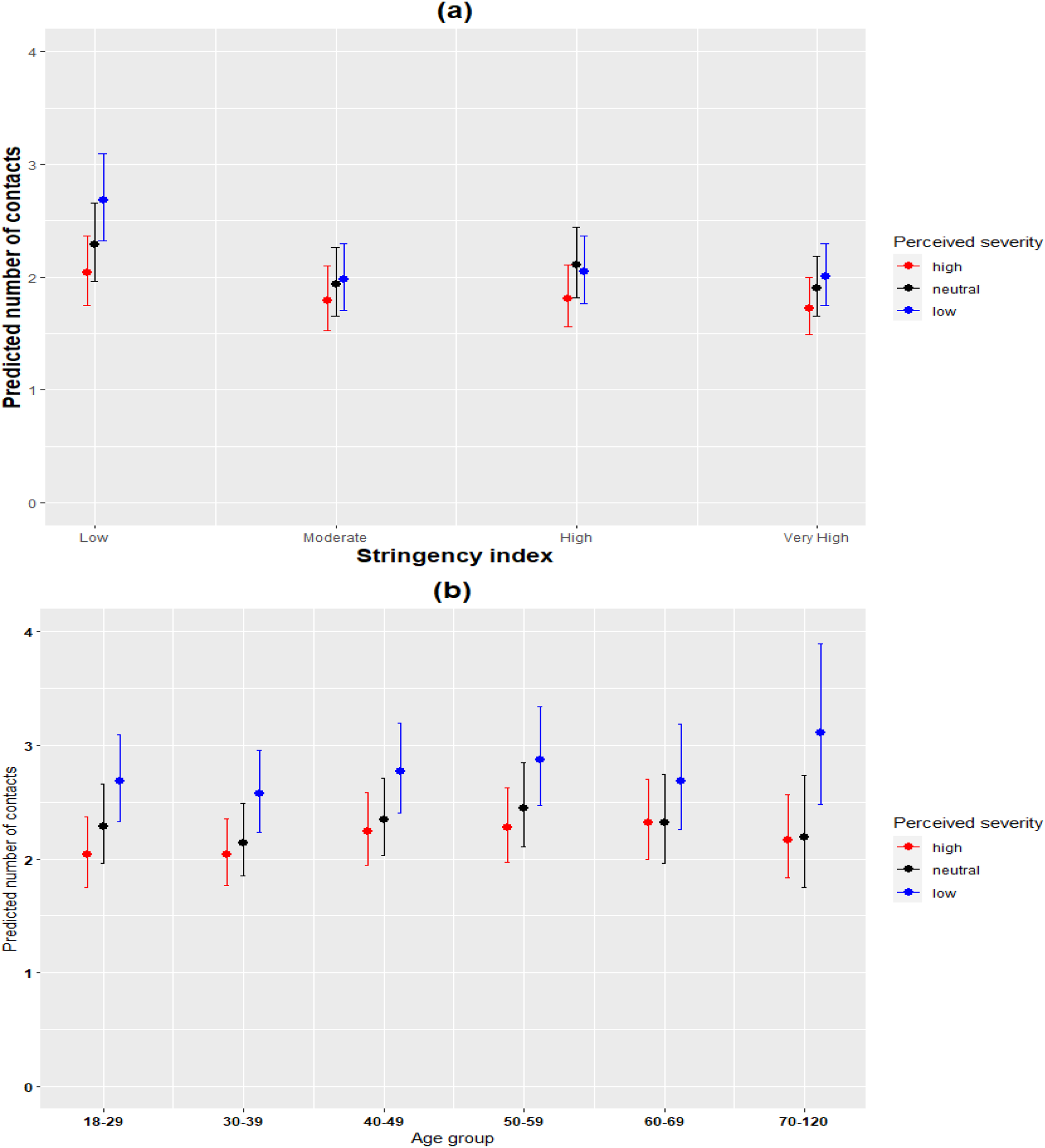
**(a):** Predicted number of contacts for perceived severity and stringency index with 95% confidence interval (CI) from the perceived severity model in the period between December 2020 and September 2021. **(b):** Predicted number of contacts for perceived severity and age group with 95% confidence interval (CI) from the perceived severity model in the period between December 2020 and September 2021.

#### Perceived susceptibility

The plots of the predicted number of contacts showed that in general, participants with high levels of perceived susceptibility reported more contacts than those with low or neutral perceptions on susceptibility (Supplementary Figure 3, Additional file 1). The results yielded a significant interaction effect between stringency index and perceived susceptibility (p-value = 0.039). See (Supplementary Table 5, Additional file 1) for the perceived susceptibility model results.

#### Perceived risk to vulnerable

The results for the perceived risk to vulnerable model did not yield insightful results. We did not find consistent patterns from the predicted number of contacts from the marginal effects of interaction between the perceived risk to the vulnerable and either vaccination status, age group, or stringency index. See (Supplementary Table 6, Additional file 1) for the model results of perceived risk to vulnerable.

#### Cross-level effects of country on social contacts

When the country was considered as a fixed effect in the model of the perceived severity, the marginal effects of the cross-level interactions between the country and perceived severity confirmed that participants with low and neutral levels of perceived severity reported more contacts than those with high levels of perceived severity (Figure 4). The interaction term between country and perceived severity was not statistically significant. The plot of the predicted number of contacts from the interaction effects between country and vaccination status further showed that vaccinated individuals reported more contacts than non-vaccinated ones in the different countries (Figure 5). Furthermore, the interaction term was statistically significant implying that the differences in the predicted mean number of contacts between the vaccinated and non-vaccinated differed in the different countries.

**Figure 4:**
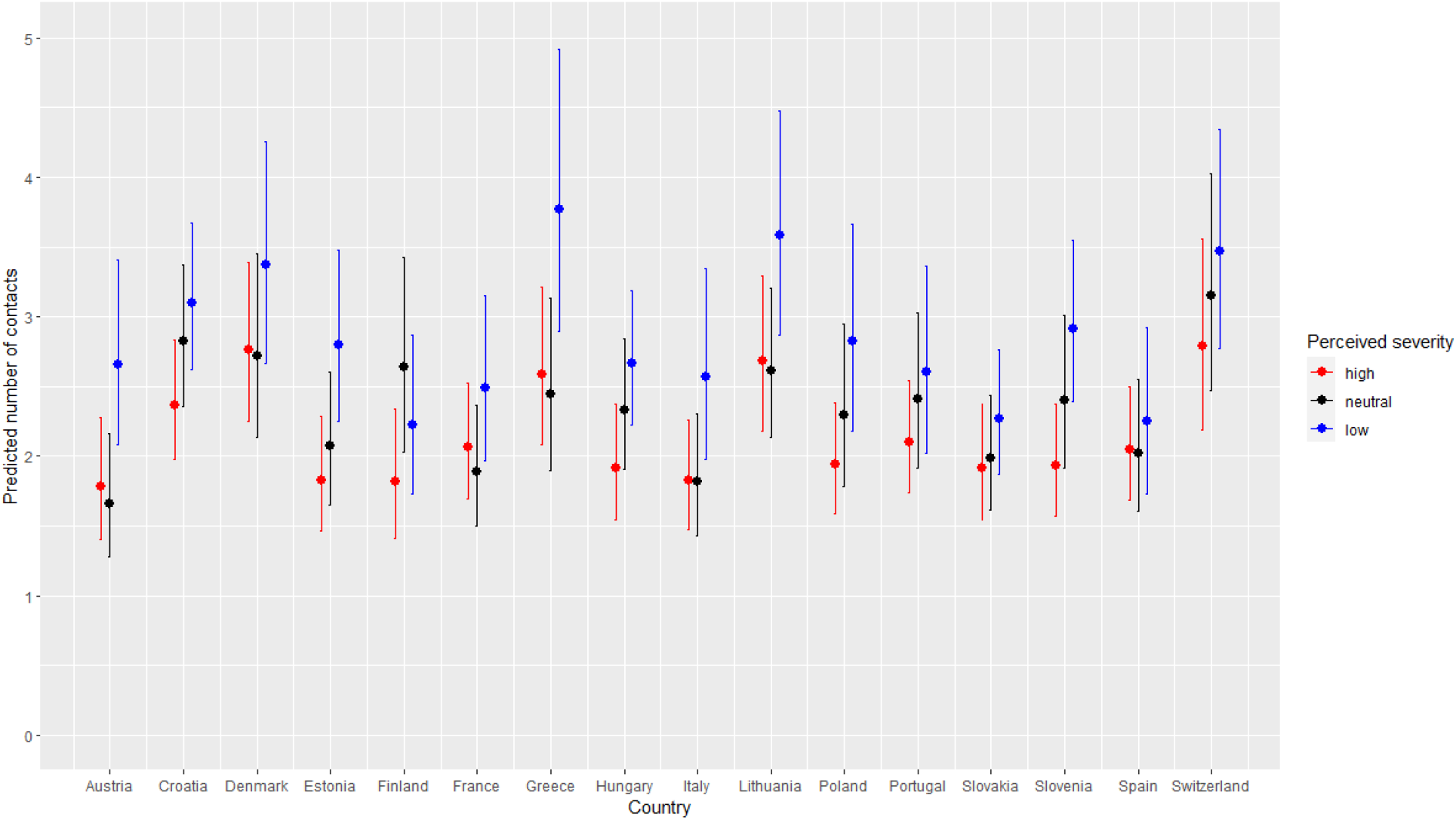
Predicted number of contacts for perceived severity and country with 95% CI from the perceived severity model with country as a fixed effect in the period between December 2020 and September 2021.

**Figure 5:**
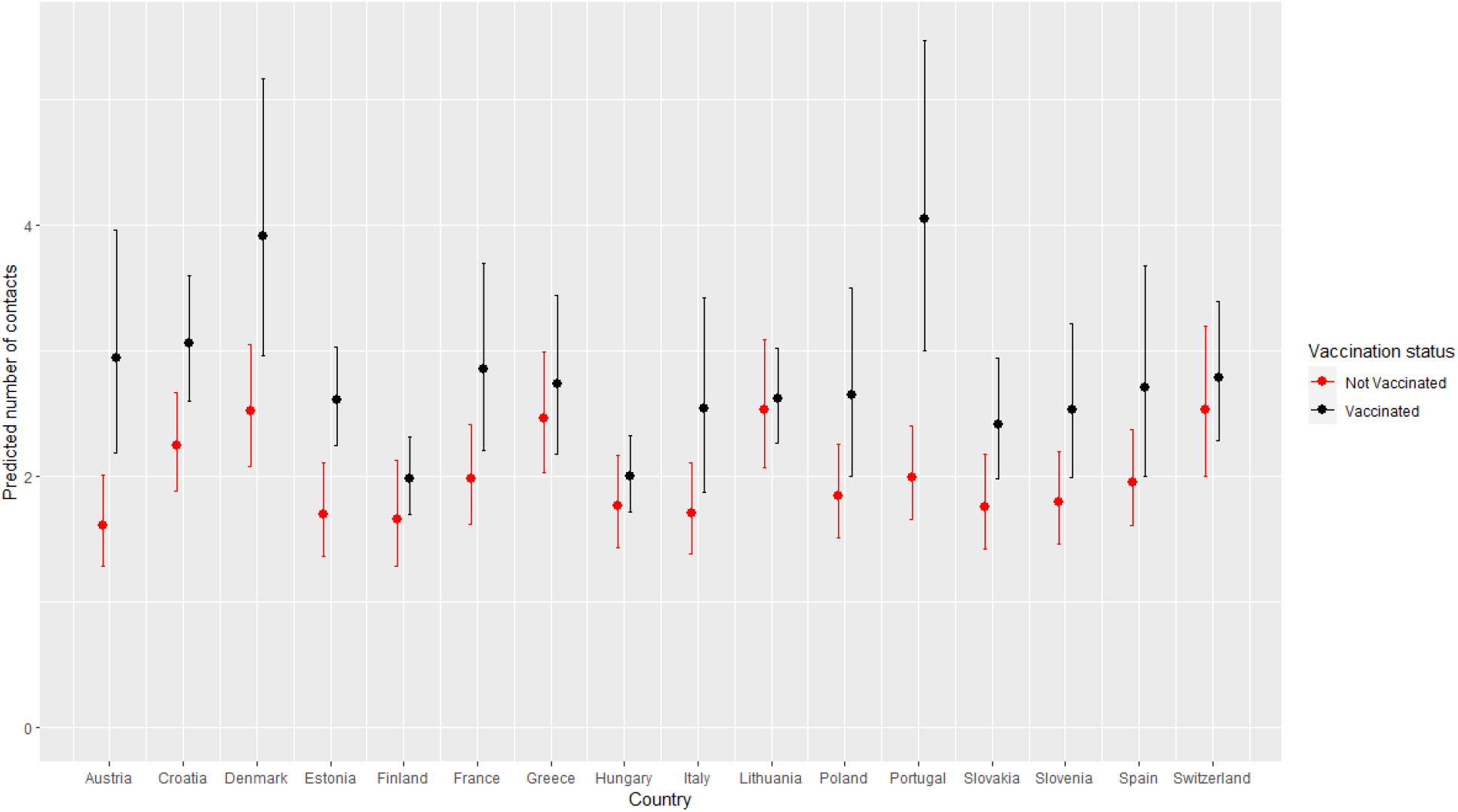
Predicted number of contacts for vaccination status and country with 95% CI from the perceived severity model with country as a fixed effect in the period between December 2020 and September 2021.

Results from the perceived susceptibility model did not show distinct patterns from the cross-level interaction effects between perceived susceptibility and country in terms of the predicted number of contacts between the participants with low, neutral, or high levels of perceived susceptibility (Supplementary Figure 4, Additional file 1). Furthermore, the cross-level interaction effects between the country and the perceived susceptibility was not statistically significant implying that there were no substantial differences in the relationship between the perceived susceptibility and number of social contacts in the different countries.

Similarly, results from the perceived risk to the vulnerable model considering country as a fixed effect indicated no significant interaction effects between perceived risk to vulnerable and country implying no major differences between countries. The predicted number of contacts did not yield significant relationships across the different levels of perceived risk to vulnerable (Supplementary Figure 5, Additional file 1).

Conducting similar analysis using the total number of contacts reported away from home yielded similar results in terms of the observed relationships in our study (Supplementary Table 7, Additional file 1). Furthermore, we performed sensitivity analyses to explore the possible influence of singletons in our analysis (i.e, participants who participated only once in the study). Excluding these singletons led to slight differences in the parameter estimates of the model. However, the observed results were robust.

Finally, we performed a preliminary analysis considering participants who indicated to have been vaccinated. Using a generalized estimating equation (GEE) model, we assessed the impact of vaccination on the individual’s risk perceptions. We found that vaccinated individuals had 1.08 (95% CI 1.02 - 1.16) and 1.17 (95% CI 1.06 - 1.28) times estimated odds for indicating low and neutral perceptions of severity, respectively, as compared to the non-vaccinated (Supplementary Table 8, Additional file 1). For the perceived susceptibility, we found that vaccinated individuals had 0.89 (95% CI 0.83 - 0.97) and 1.70 (95% CI 1.56 - 1.85) times estimated odds for indicating low and neutral perceptions of susceptibility, respectively, as compared to the non-vaccinated. Whilst for the risk to vulnerable, vaccinated individuals had 0.88 (95% CI 0.86 - 0.96) and 1.22 (95% CI 1.09 - 1.35) times estimated odds of indicating low and neutral perceptions of risk to vulnerable, respectively, as compared to the non-vaccinated. Given the importance of vaccine-induced behavioral changes and risk perception, future research should focus on characterizing their relation, especially in relation with social contact data.

## Discussion

The main objective of this study was to explore the influence of COVID-19 vaccination status and the COVID-19 related risk perceptions on the number of social contacts. We analyzed longitudinal data collected in 16 European countries using a multilevel generalized linear mixed effects model to account for within and between participants variations while controlling for the hierarchical structure of the data. Furthermore, we also performed cross-level analysis to explore the relationships between both the perceptions and vaccination status with number of social contacts in the different countries.

The results indicated that perceived severity and vaccination status played a crucial role in modulating the number of social contacts. More specifically, we found that individuals who had high levels of perceived severity reported fewer social contacts as compared to those who had low and neutral levels of perceived severity. The observed associations are consistent with the results found in analysis of CoMix data limited for Belgium [26], and UK [29]. The Belgian CoMix study encompassed two longitudinal surveys, one between April 2020 and August 2020, and the other between November 2020 and April 2021. The results indicated that in the first survey, participants who had low and neutral levels of perceived severity reported 70% and 56% more contacts as compared to those who had high perception of severity. Whilst in the second survey, the participants with low and neutral perceptions on perceived severity reported 62% and 76% more contacts than those who had high perceived severity. The UK CoMix study [29] on the other hand utilized data collected between March 2020 and March 2021. Applying clustered bootstrapping to obtain the mean number of social contacts, the results indicated that participants who had low levels of perceived severity reported more contacts than those who had high levels of perceived severity. The similarity in the observed relationships in these studies further highlights the crucial role of perceptions in modulating social contacts.

The predicted number of contacts in individuals who were not vaccinated against the SARS-CoV-2 were lower than individuals who had received a vaccine. These differences were consistent in all the countries included in this study and is also consistent with a recent study that found that vaccinated individuals generally had more contacts than the unvaccinated [43]. This implies that vaccination against COVID-19 played a crucial role in shaping the social contact behaviour during the COVID-19 crisis. This could be due, among other factors, to the minor restrictions that vaccinated individuals were subjected to in several European countries, where the vaccination certificate was one of the necessary condition to access public or indoor areas. In such a circumstance, the increase in contacts could be related also to the higher potential of social interactions of vaccinated individuals. However, since our analysis showed that individuals change their risk perceptions after vaccination, the increase in contacts is most likely due to a combination of a higher potential for social interactions and a spontaneous behavioral change. These apparent changes in behaviour following COVID-19 vaccination could have important implications in the context of the COVID-19 pandemic which has been characterized by the emergence of variants of concern such as Omicron and its sub-variants [13, 14]. These mutations initiated uncertainties in the effectiveness of vaccines in conferring protection to the vaccinated [30]. Thus, an increase in social contact behaviour following vaccination could be disastrous for viral transmission dynamics in the event of emergence of an immune escaping variant.

Our results also indicated relatively little variation in social contact behaviour between countries, once all other confounders are accounted for. This low variation could be a result of the stringent measures implemented to limit the number of social contacts during the different phases of COVID-19 pandemic in the different countries. Interestingly, we observed substantial heterogeneity in social contact behaviour between individuals. This suggests that individual-level factors played a more substantial role in influencing contact behaviour than country-level factors. The underlying heterogeneity in social contact behaviour is consistent with results from social contact studies conducted before and during the COVID-19 pandemic [44, 45].

In our study, the relationship between perceived susceptibility and perceived risk to vulnerable with the number of contacts did not yield similar patterns across different countries. This warrants more research in order to gain insights on their relevance in influencing social contacts in the context of COVID-19. From the analyses of the perceptions, we found that perceptions of susceptibility to infections were in general the lowest in comparison with perceptions on severity and risk to the vulnerable. This is indicative of the possible presence of optimism bias, a situation characterized by individuals tending to under-estimate the probability of acquiring infections in the context of infectious disease epidemiology [46]. The lack of significant relationships between the perceived risk to vulnerable could be a result of the subjective nature of perceived risk to others. Participants who do not have vulnerable individuals in their social networks might perceive low risk as compared to those that usually interact with vulnerable individuals. A study performed during the COVID-19 revealed that individuals perceive different risks for COVID-19 on their own health as compared to others such as family, friends and the general community [47]. Another study showed that individuals who were more concerned about spreading COVID-19 to vulnerable people made more contacts at home [48]. Thus the lack of association between the perceived risk to the vulnerable and social contact behavior could suggest a more diverse meaning for this risk perception construct by different people given different experiences and interactions with vulnerable people in the community.

Our study explicitly used the number of social contacts as the response variable and considered three separate constructs related to the risk perceptions. Other studies have shown that COVID-19 risk perceptions play a crucial role on adoption of protective health behaviours [20, 21, 22, 23, 24, 25, 26]. Although most of these studies are conducted in individual countries, a recent multi-country cross-sectional study by Dryhurst et al [23] conducted in ten countries across Europe, Asia and America found similar associations between risk perception and the adoption of the protective health behaviours. It is worthy mentioning that most studies exploring the influence of risk perceptions on adoption of protective behaviours use cross-sectional data yielding insights on only one time point and thus the dynamical aspects of the changing pandemic situation is not taken into account. Conversely, a recent study found that risk perceptions played a significant role on the adoption of recommended health behaviours over time in the UK [24]. Thus a novelty of our study, that employed multi-country longitudinal data, was to confirm such a relation in an evolving pandemic situation and for different countries. Furthermore, as social contacts can be used to inform models of infectious diseases, our results on risk perceptions and behavioural changes following vaccination can be incorporated in future mathematical models for a more granular dynamical exploration during a pandemic.

Our work is subjected to several limitations. The reporting of the number of social contacts was done retrospectively, hence could suffer from recall bias. However, such an effect is expected to be small since participants reported contacts in the day preceding the survey day. Due to the longitudinal nature of data collection, participants could experience response fatigue posing concerns on the quality of the data collected. Assessing the possible presence and subsequent influence of response fatigue will be studied in future. This study utilized a multi-level generalized linear mixed effects model. However, there have been concerns about the appropriateness of multi-level models when utilizing multi-country data where the number of the countries is relatively small (i.e, 25 for linear models and 30 for logit models) and the number of individuals per country is large [49]. We expect a small impact on the reliability of the estimates of our individual level effects, as the relatively small number of countries only affects the estimates of the country level predictors. The study only relied on the Oxford stringency index which gives varying weights to the diverse NPIs and collapses them into a single composite index. Future work can compare analyses using different NPIs databases, including the Response Measures Database by the European Centre for Disease Prevention and Control (ECDC) and the European Commission’s Joint Research Centre [50]. Lastly, it is crucial to mention that although the surveys in each individual country were representative in terms of age, gender, and also region of residence in the panel of participants considered, the optional participation in each subsequent round of data collection could suffer from self-selection bias. The average participation was generally high in all the countries.

## Conclusion

In this study, we utilized longitudinal data from a panel of individuals from 16 European countries collected between December 2020 and September 2021 to explore the influence of COVID-19 vaccination status and related COVID-19 risk perceptions on the number of social contacts. We found little differences in social contact behaviour between the countries. However, there were marked heterogeneity in individual social contact behaviour. We found that individuals who had high levels of perceived severity of COVID-19 reported significantly fewer number of social contacts in comparison with those who had low or neutral levels of perceived severity. Furthermore, vaccinated individuals reported significantly more contacts than the non-vaccinated. Thus our study adds important insights into the significance of perceived severity on social contact behaviour from a multi-country perspective. Further, it highlights the subsequent changes in social contact behaviour following vaccination. This could be potentially disastrous if appropriate action is not taken in the event of the emergence of an immune escaping variant, since vaccination in that situation would lead to an increase in contacts but not an advantage in terms of protection, resulting in a higher disease burden. These considerations should be taken into account when designing, implementing and communicating COVID-19 interventions. Owing to the importance of social contact behaviour in the transmission dynamics of infectious diseases, further research is needed to disentangle the relation between contacts, vaccination and perception.

## Data Availability

The datasets utilized in this study are available in the Zenodo-based repository,
www.socialcontactdata.org/data

https://www.bsg.ox.ac.uk/research/publications/variation-government-responses-covid-19

## List of abbreviations

ECDC: European Centre for Disease Prevention and Control
HERA: Health Emergency Preparedness and Response Authority
NPI: Non-Pharmaceutical Intervention
HBM: Health Belief Model
PMS: Protection Motivation and Self-efficacy
UK: United Kingdom
OxCGRT: Oxford COVID-19 Government Response Tracker
GLMM: Generalized Linear Mixed effects Model
SD: Standard Deviation
CI: Confidence Interval
GEE: Generalized Estimating Equations NA : Not Applicable

## Declarations

### Ethics approval and consent to participate

Approvals or waivers for the CoMix study questionnaires and protocols were obtained through the local ethical committees for each individual country in the study. More details can be found in Verelst et al [33].

### Consent for publication

Not applicable

### Availability of data and materials

The datasets utilized in this study are available in the Zenodo-based repository, www.socialcontactdata.org/data

### Authors’ contributions

JW, PC, HJ and NH conceptualised the study. CIJ and WJE designed the initial CoMix survey in consultation with PB and NH. KW, PC and JW cleaned the data. JW analysed the data with all authors contributing in the analysis and interpretation of results. JW wrote the first draft. All authors contributed and reviewed the manuscript and approved the final version for publication.

## Acknowledgements

Not applicable.

