## Supplementary Information for "The influence of COVID-19 risk perception and vaccination status on the number of social contacts across Europe: insights from the CoMix study"

November 23, 2022

[1]Data Science Institute, I-BioStat, Hasselt University, Hasselt, Belgium.

[2]Centre for Mathematical Modelling of Infectious Diseases, Department of Infectious Disease Epidemiology, London School of Hygiene and Tropical Medicine, Keppel Street, WC1E 7HT London, UK.

[3]European Centre for Disease Prevention and Control (ECDC), Gustav III:s Boulevard 40, 169 73 Solna, Sweden.

[4]Centre for Health Economics Research and Modelling Infectious Diseases, Vaccine & Infectious Disease Institute, University of Antwerp, Antwerp, Belgium.

[5]The University of New South Wales, School of Public Health and Community Medicine, Sydney, NSW 2033, Australia.

[**] Current address: Health Emergency Preparedness and Response Authority (HERA), European Commission, 1049 Brussels, Belgium.

### Supporting information

| Country | Start date of data collection | End date of data collection | Total survey rounds in Adults | Total survey rounds in Children |
|---|---|---|---|---|
| Austria | 22 December 2020 | 19 April 2021 | 7 | 2 |
| Denmark | 22 December 2020 | 16 April 2021 | 7 | 2 |
| France | 21 December 2020 | 08 April 2021 | 7 | 2 |
| Italy | 21 December 2020 | 23 April 2021 | 7 | 2 |
| Portugal | 22 December 2020 | 22 April 2021 | 7 | 2 |
| Poland | 22 December 2020 | 19 April 2021 | 7 | 2 |
| Spain | 21 December 2020 | 12 May 2021 | 7 | 2 |
| Greece | 18 February 2021 | 08 June 2021 | 7 | 2 |
| Slovenia | 04 March 2021 | 09 June 2021 | 7 | 2 |
| Croatia | 20 April 2021 | 18 August 2021 | 7 | 2 |
| Estonia | 20 April 2021 | 23 August 2021 | 7 | 2 |
| Hungary | 20 April 2021 | 18 August 2021 | 6 | 2 |
| Slovakia | 20 April 2021 | 16 August 2021 | 7 | 2 |
| Switzerland | 3 June 2021 | 15 September 2021 | 6 | 2 |
| Lithuania | 2 July 2021 | 30 September 2021 | 6 | 2 |
| Finland | 2 July 2021 | 30 September 2021 | 6 | 2 |

Supplementary Table 1: Country specific timelines of the CoMix data collection in the 16 countries included in the study.

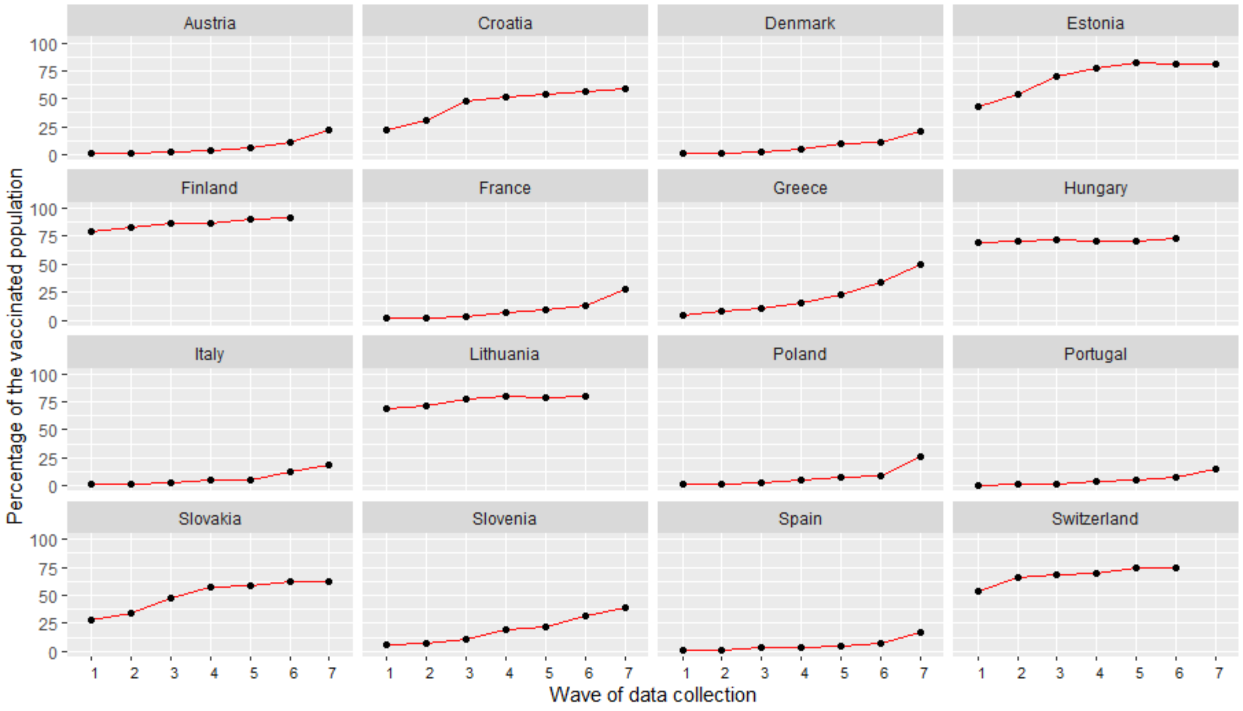


Supplementary Figure 1: Percentage of the adult population (18 years or above) vaccinated with the first dose of any of the COVID-19 vaccine during the data collection period in the different countries.

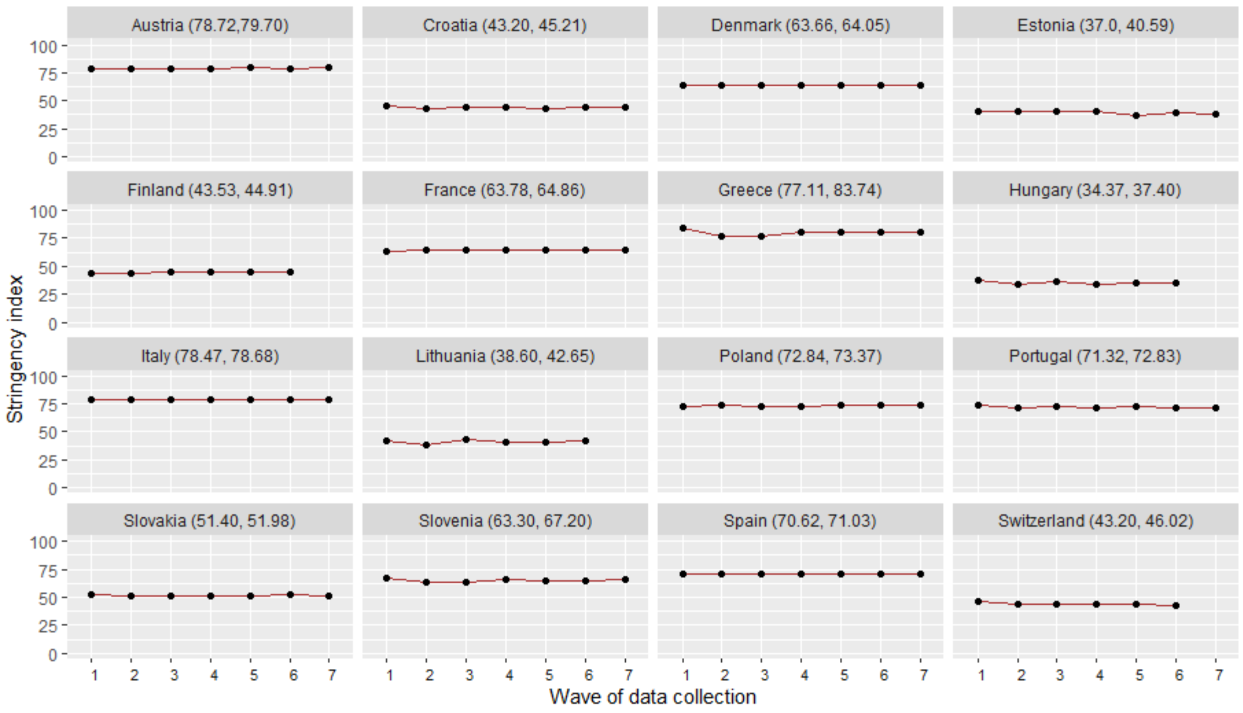


Supplementary Figure 2: Variations in the stringency index during the data collection period in the different countries.

### 0.1 Reliability analysis for internal consistency

The risk perceptions consisted of three statements that the participants were asked to respond to. The first statement was *"I am likely to catch coronavirus"*, the second was *"I am worried that I might spread coronavirus to someone who is vulnerable"*, and the third was *"coronavirus would be a serious illness for me"*. These items were coded on 5-point Likert scales. The response option *"do not know"* was treated as a missing value in the reliability analysis, but was included in the modeling analysis as explained below.

We utilized Cronbach's alpha as the reliability measure of internal consistency of the Likert-scales of the risk perception items [1]. We considered a threshold of 0.80 of the Cronbach's alpha reliability coefficient to group the items [1]. The three risk perceptions items had an overall Cronbach's alpha of 0.670 (95% confidence interval (CI) 0.669 - 0.671) for all the countries combined. The Cronbach's alpha for each individual country ranged between 0.33 (95% CI 0.30 - 0.36) and 0.76 (95% CI 0.75 - 0.77). More information on the Cronbach's alpha for each country is detailed in Supplementary Table 11. Thus, following the resulting overall Cronbach's alpha for the three items which was below the considered threshold, we did not combine the items into one composite variable to represent the underlying risk perception construct. Therefore, the three risk perception items were considered to measure three separate constructs. We refer to these constructs as, *perceived severity* for the item *"coronavirus would be a serious illness for me"*, *perceived susceptibility* for the item *"I am likely to catch coronavirus"*, and *perceived risk to the vulnerable* for the item *"I am worried that I might spread coronavirus to someone who is vulnerable"*. Motivated by exploratory analyses, we categorised the 6 response options into three response levels: low perception ('tend to disagree' and 'strongly disagree'), neutral ('neither agree nor disagree' and 'don't know') and high perception ('strongly agree' and 'tend to agree').

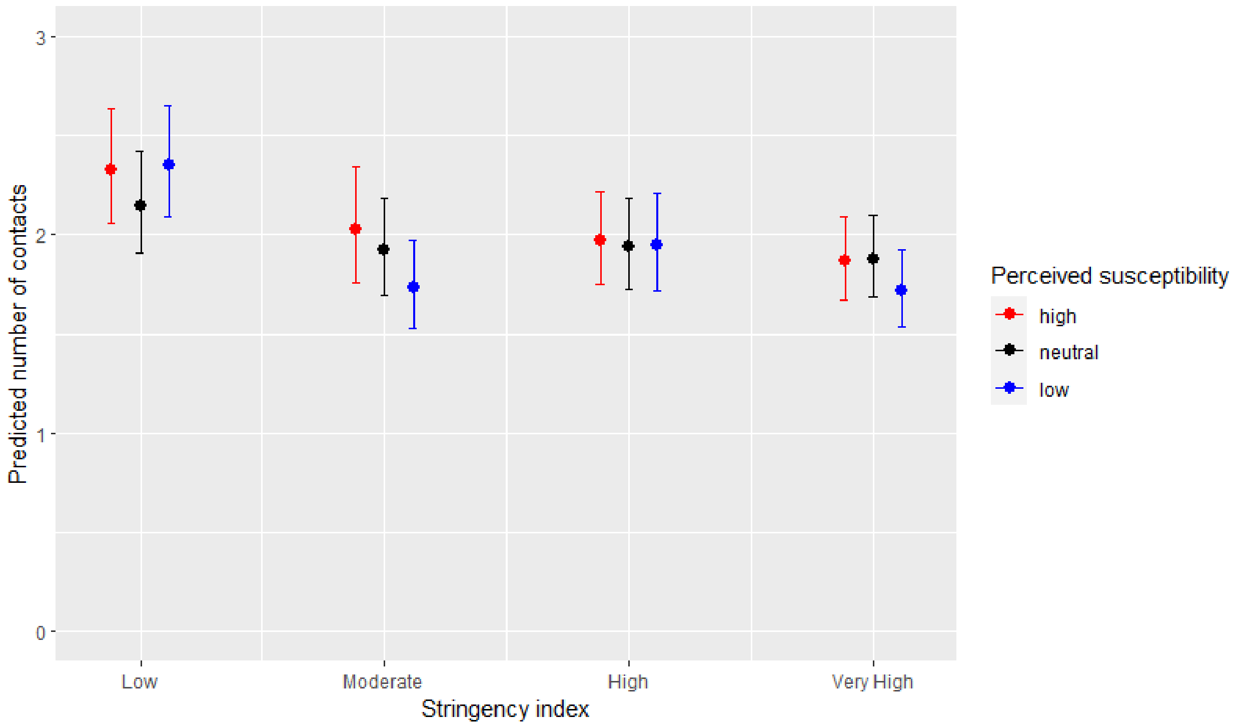


Supplementary Figure 3: Predicted number of contacts by perceived susceptibility and stringency index with 95% CI for the perceived susceptibility model in the period between December 2020 and September 2021.

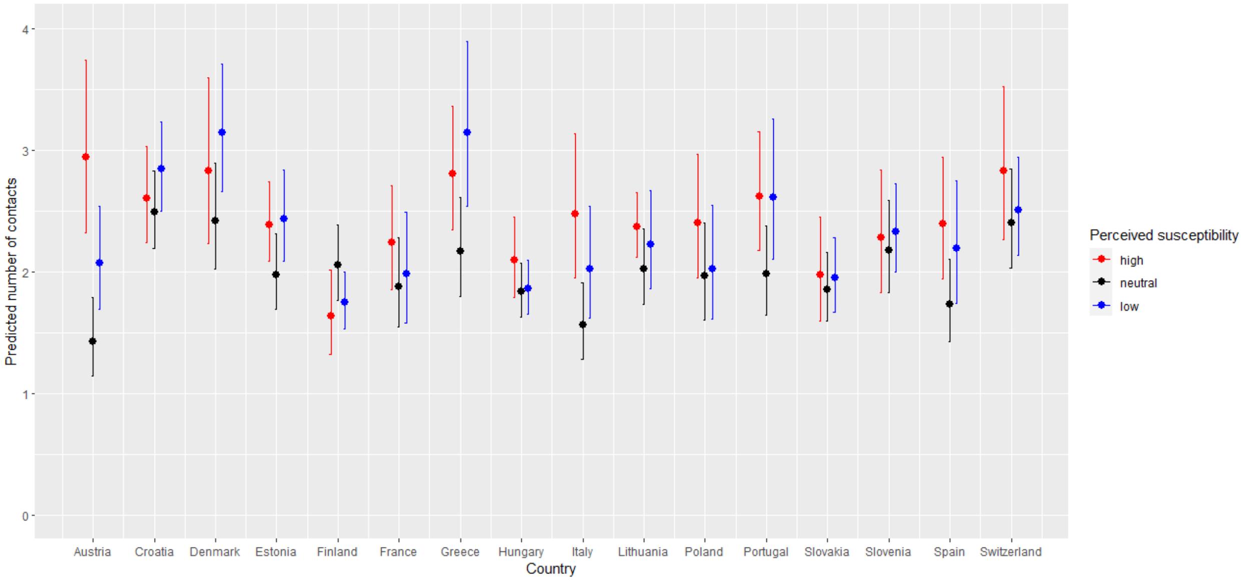


Supplementary Figure 4: Predicted number of contacts from the marginal effects of interaction between perceived susceptibility and country with 95% CI for the perceived susceptibility model with country as a fixed effect in the period between December 2020 and September 2021.

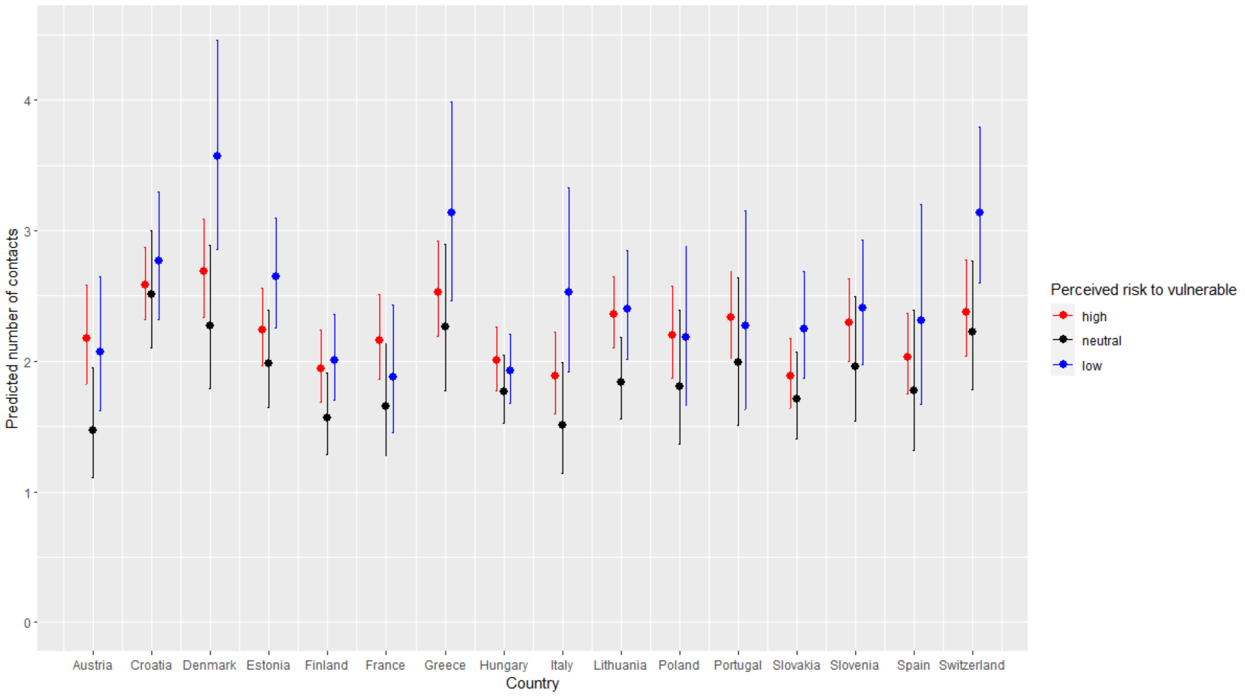


Supplementary Figure 5: Predicted number of contacts from the marginal effects of interaction between perceived risk to vulnerable and country with 95% CI for the perceived risk to vulnerable model with country as a fixed effect in the period between December 2020 and September 2021.

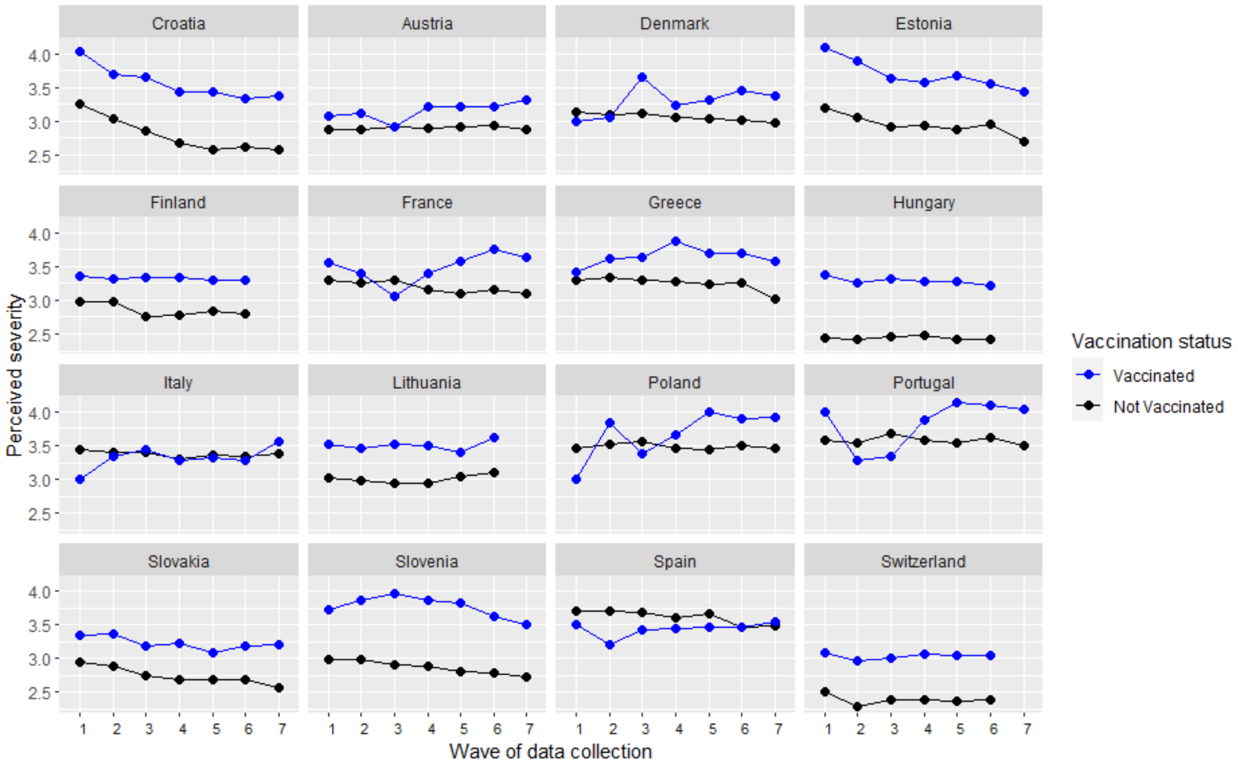


Supplementary Figure 6: Mean score of the perceived severity during the data collection period in the different countries.

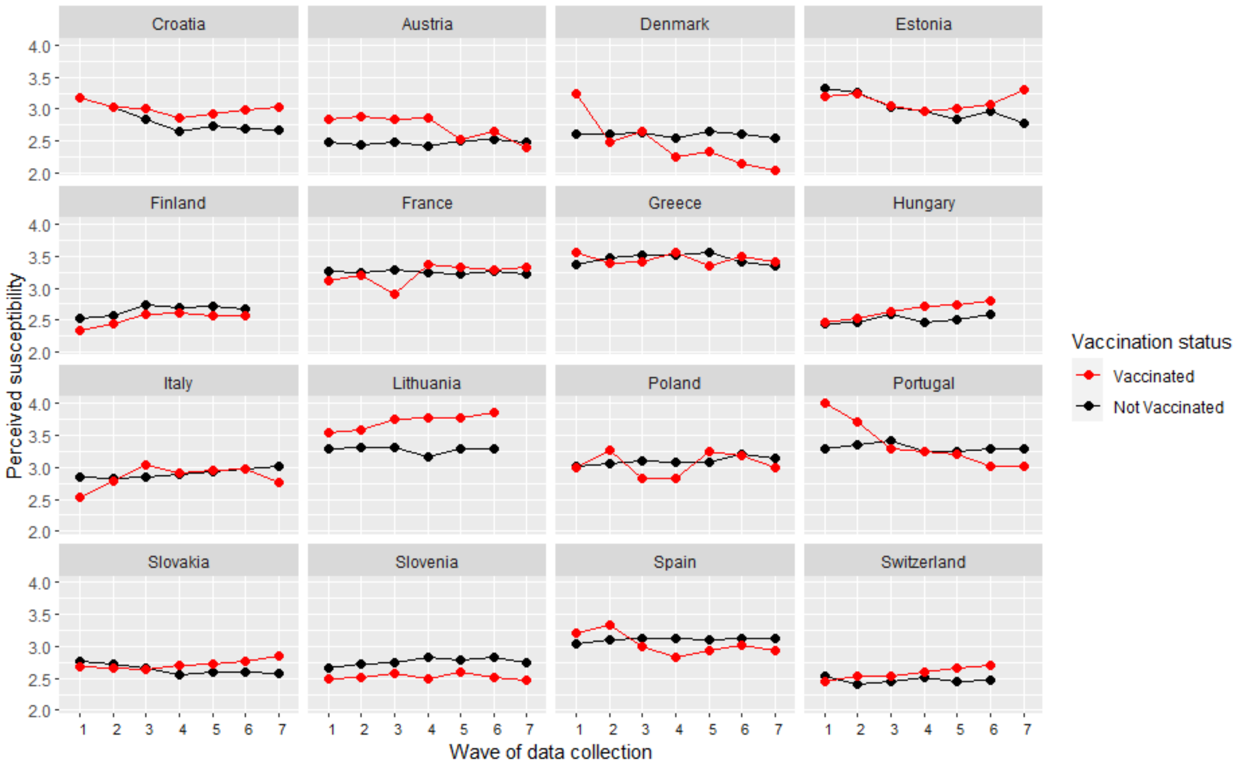


Supplementary Figure 7: Mean score of the perceived susceptibility during the data collection period in the different countries.

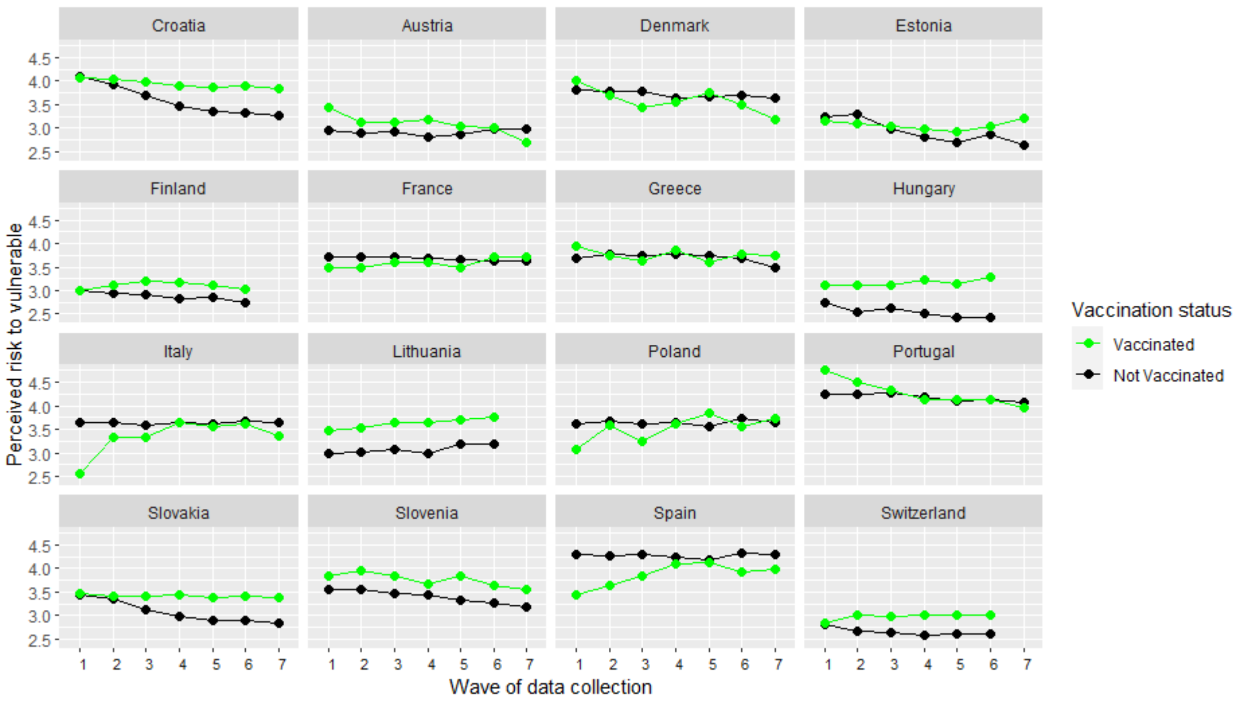


Supplementary Figure 8: Mean score of the perceived risk to the vulnerable during the data collection period in the different countries.

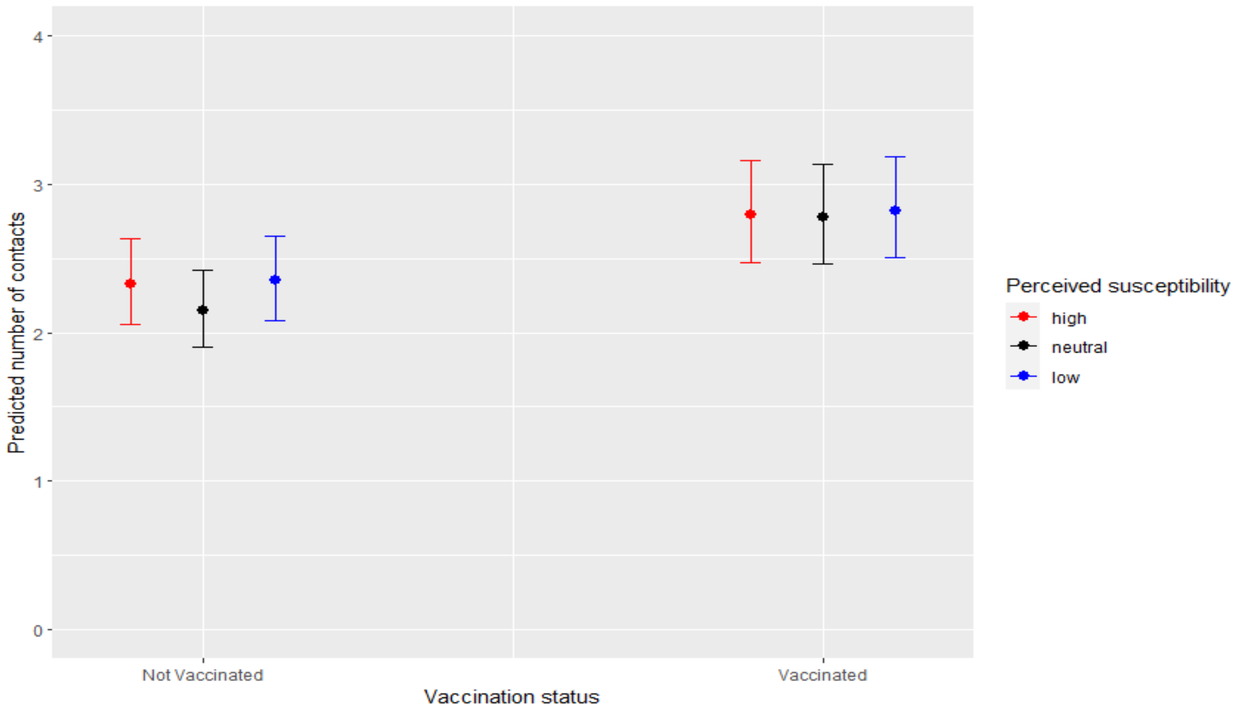


Supplementary Figure 9: Predicted number of contacts by perceived susceptibility and vaccination status with 95% CI for the perceived risk to vulnerable model in the period between December 2020 and September 2021.

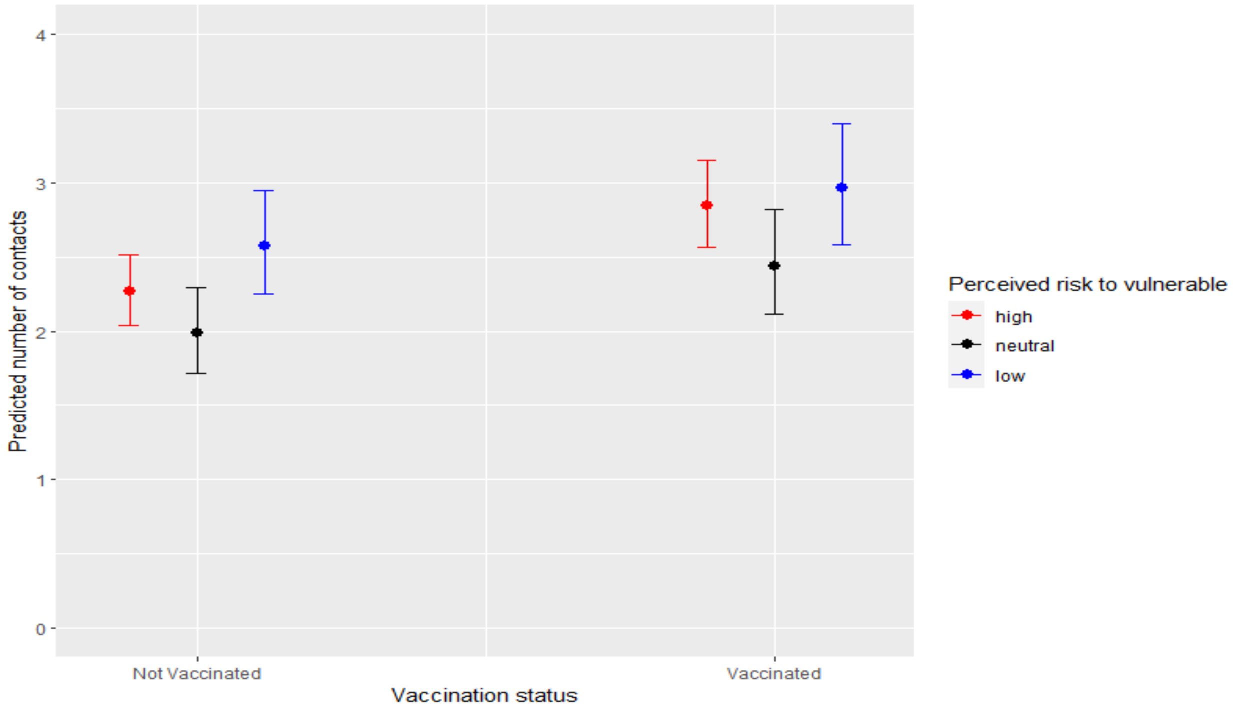


Supplementary Figure 10: Predicted number of contacts by perceived risk to vulnerable and vaccination status with 95% CI for the perceived risk to vulnerable model in the period between December 2020 and September 2021.

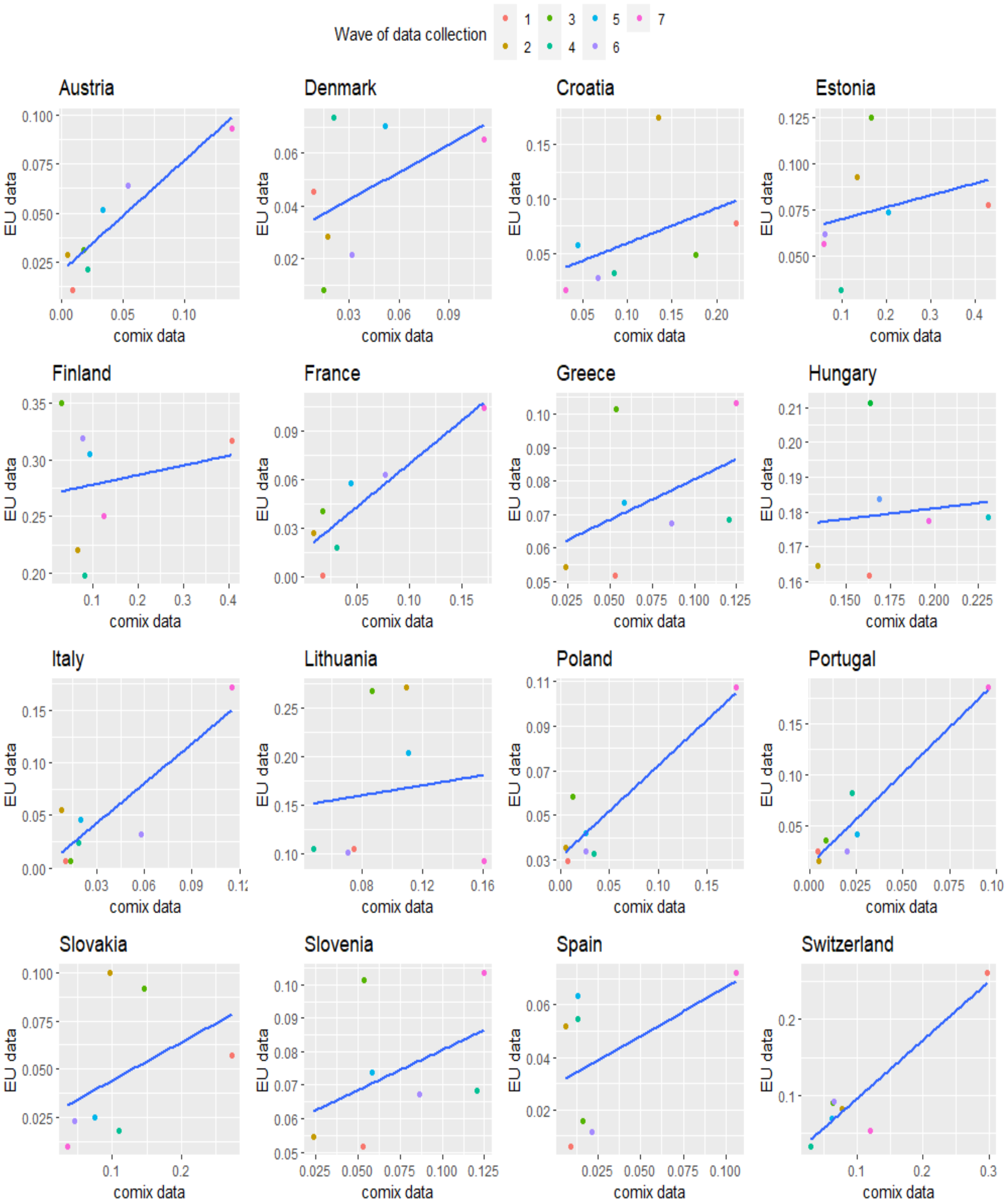


Supplementary Figure 11: Comparison of the percentages of the vaccinated between the CoMix data and the ECDC data.

| Country | Total participants | Total surveys | Gender Female (%) | Gender Male (%) | Age group 18-29 (%) | Age group 30-39 (%) | Age group 40-49 (%) | Age group 50-59 (%) | Age group 60-69 (%) | Age group 70-120 (%) | Average survey comp. |
|---|---|---|---|---|---|---|---|---|---|---|---|
| Austria | 1738 | 7642 | 48.4 | 51.4 | 18.5 | 17.3 | 16.8 | 19.5 | 18.5 | 9.4 | 4.05 |
| Croatia | 1971 | 7080 | 51.4 | 48.6 | 18.4 | 16.4 | 20.0 | 18.1 | 21.6 | 5.4 | 3.59 |
| Denmark | 1730 | 7057 | 47.1 | 52.7 | 14.1 | 10.6 | 13.7 | 22.4 | 22.8 | 16.4 | 4.07 |
| Estonia | 1857 | 6986 | 54.0 | 45.7 | 13.2 | 17.0 | 16.6 | 18.3 | 18.6 | 16.3 | 3.76 |
| Finland | 1620 | 6444 | 48.0 | 51.6 | 14.8 | 15.4 | 15.2 | 17.0 | 23.4 | 14.3 | 3.97 |
| France | 1710 | 7047 | 46.4 | 53.2 | 15.0 | 17.4 | 16.5 | 15.3 | 22.4 | 13.4 | 4.12 |
| Greece | 2245 | 7130 | 48.5 | 51.2 | 16.1 | 21.2 | 26.3 | 24.4 | 10.0 | 2.0 | 3.18 |
| Hungary | 2085 | 6421 | 47.4 | 52.3 | 12.9 | 17.1 | 22.1 | 16.4 | 22.2 | 9.4 | 3.07 |
| Italy | 1617 | 7054 | 46.6 | 53.3 | 15.0 | 13.5 | 21.8 | 15.7 | 20.5 | 13.4 | 4.36 |
| Lithuania | 1918 | 6429 | 54.8 | 45.2 | 14.3 | 17.8 | 16.6 | 19.1 | 22.4 | 9.8 | 3.35 |
| Poland | 1724 | 7011 | 55.6 | 44.4 | 18.8 | 19.3 | 19.2 | 18.0 | 18.9 | 5.8 | 4.06 |
| Portugal | 1840 | 7031 | 45.5 | 54.3 | 16.0 | 16.1 | 21.8 | 19.2 | 18.4 | 8.5 | 3.82 |
| Slovakia | 1733 | 7049 | 47.4 | 52.6 | 15.3 | 20.5 | 20.5 | 16.5 | 19.9 | 7.2 | 4.07 |
| Slovenia | 1998 | 7510 | 56.7 | 43.2 | 15.8 | 20.3 | 21.4 | 20.9 | 18.1 | 3.5 | 3.76 |
| Spain | 1622 | 7059 | 49.5 | 50.3 | 14.6 | 16.9 | 22.4 | 17.7 | 18.8 | 9.4 | 4.35 |
| Switzerland | 1884 | 6758 | 48.7 | 50.9 | 15.4 | 18.4 | 16.3 | 20.8 | 17.2 | 11.9 | 3.59 |

Supplementary Table 2: Summary of the sample characteristics of the countries included in the analysis.

| Perceived severity Model | | | |
|---|---|---|---|
| Predictors | Incidence rate ratios | Confidence intervals (CI) | P-value |
| household size | 1.29** | 1.28 - 1.30 | < 0.001 |
| Gender[Female]-Ref | - | - | - |
| Gender[Male] | 0.90** | 0.88 - 0.93 | <0.001 |
| Employment status [full time] - Ref | - | - | - |
| Employment status [part-time (<34 hours)] | 0.96 | 0.90 - 1.02 | 0.202 |
| Employment status [full time homemaker] | 0.79** | 0.73 -0.86 | <0.001 |
| Employment status [long term sick or disabled] | 0.72** | 0.63 - 0.82 | <0.001 |
| Employment status [Retired] | 0.70** | 0.64 - 0.74 | <0.001 |
| Employment status [Self employed] | 0.96 | 0.90 - 1.03 | 0.266 |
| Employment status [Student/pupil] | 0.88 | 0.82 - 0.93 | <0.001 |
| Employment status [Unemployed and not looking for a job] | 0.72** | 0.63 - 0.83 | <0.001 |
| Employment status [Unemployed and looking for a job] | 0.71** | 0.67 - 0.75 | <0.001 |
| High risk status [No] - Ref | | | |
| High risk status [Yes] | 0.98 | 0.94 - 1.02 | 0.289 |
| History of infection [Infected] - Ref | | | |
| History of infection [Not infected] | 1.01 | 0.93 - 1.09 | 0.852 |
| History of infection [Not tested] | 0.92 | 0.85 - 0.99 | 0.026 |
| Vaccination status [Not vaccinated] - Ref | | | |
| Vaccination status [vaccinated] | 1.31** | 1.23 - 1.39 | <0.001 |
| Perceived severity [high perception] - Ref | - | - | - |
| Perceived severity [low perception] | 1.25** | 1.13 - 1.37 | <0.001 |
| Perceived severity [neutral perception] | 1.10 | 1.0 - 1.21 | 0.091 |
| Day of week[weekday] - Ref | | | |
| Day of week[weekend] | 0.90 | 0.86 - 0.93 | <0.001 |
| Age group [18-29] - Ref | - | - | - |
| Age group [30-39] | 0.95 | 0.90 - 1.00 | 0.072 |
| Age group [40-49] | 1.02 | 0.97 - 1.08 | 0.424 |
| Age group [50-59] | 1.04 | 0.98 - 1.10 | 0.117 |
| Age group [60-69] | 1.06 | 0.98 - 0.113 | 0.129 |
| Age group [70-120] | 1.01 | 0.91 - 1.11 | 0.772 |
| Stringency index [low] -Ref | - | - | - |
| Stringency index [Moderate] | 0.92 | 0.85 - 0.95 | 0.001 |
| Stringency index [High] | 0.87** | 0.80 - 0.95 | 0.001 |
| Stringency index [Very high] | 0.84** | 0.77 - 0.92 | <0.001 |
| Vaccination status* Perceived severity [low perception] | 0.88** | 0.80 - 0.96 | <0.001 |
| Vaccination status* Perceived severity [neutral perception] | 0.94 | 0.86 - 1.02 | 0.152 |
| Age group [18-29]* Perceived severity [low perception] -Ref | - | - | - |
| Age group [30-39]* Perceived severity [low perception] | 0.95 | 0.82 - 1.09 | 0.462 |
| Age group [40-49]* Perceived severity [low perception] | 0.92 | 0.80 - 1.05 | 0.213 |
| Age group [50-59]* Perceived severity [low perception] | 0.95 | 0.82 - 1.10 | 0.496 |
| Age group [60-69]* Perceived severity [low perception] | 0.86 | 0.74 - 1.01 | 0.065 |
| Age group [70-120]* Perceived severity [low perception] | 1.08 | 0.87 - 1.33 | 0.491 |
| Age group [18-29]* Perceived severity [neutral perception] -Ref | - | - | - |
| Age group [30-39]* Perceived severity [neutral perception] | 0.92 | 0.79 - 1.07 | 0.293 |
| Age group [40-49]* Perceived severity [neutral perception] | 0.92 | 0.80 - 1.06 | 0.267 |
| Age group [50-59]* Perceived severity [neutral perception] | 0.97 | 0.83 - 1.12 | 0.652 |
| Age group [60-69]* Perceived severity [neutral perception] | 0.88 | 0.75 - 1.03 | 0.111 |
| Age group [70-120]* Perceived severity [neutral perception] | 0.90 | 0.73 -1.11 | 0.313 |
| Stringency index [low]* Perceived severity [low perception] -Ref | - | - | - |
| Stringency index [Moderate]* Perceived severity [low perception] | 0.85 | 0.75 - 0.96 | 0.008 |
| Stringency index [High]* Perceived severity [low perception] | 0.88 | 0.78 - 1.00 | 0.050 |
| Stringency index [Very high]* Perceived severity [low perception] | 0.93 | 0.84 - 1.04 | 0.217 |
| Stringency index [low]* Perceived severity [neutral perception] -Ref | - | - | - |
| Stringency index [Moderate]* Perceived severity [neutral perception] | 1.00 | 0.89 - 1.12 | 0.982 |
| Stringency index [High]* Perceived severity [neutral perception] | 1.02 | 0.91 - 1.14 | 0.781 |
| Stringency index [Very high]* Perceived severity [neutral perception] | 0.98 | 0.88 - 1.10 | 0.723 |

Supplementary Table 3: Results from the hierarchical generalized linear mixed effects model showing the incidence rate ratios, the associated confidence intervals and p-values for the perceived severity model.

| Perceived severity Model | | | |
|---|---|---|---|
| Variable | Chisq | Df | Pr (> chisq) |
| Household size | 1553.604 | 1 | < 0.001 |
| Gender | 43.524 | 1 | <0.001 |
| Employment status | 260.414 | 8 | < 0.001 |
| High risk status | 1.122 | 1 | 0.355 |
| Stringency index | 15.463 | 3 | 0.001 |
| History of infection | 28.794 | 2 | 0.001 |
| Age group | 17.634 | 5 | 0.003 |
| Perceived severity | 21.369 | 2 | <0.001 |
| Vaccination status | 76.989 | 1 | <0.001 |
| Day of week | 33.397 | 1 | <0.001 |
| Stringency index : Perceived severity | 8.035 | 6 | 0.063 |
| Vaccination status : Perceived severity | 8.881 | 2 | 0.011 |

Supplementary Table 4: Analysis of Deviance (Type III Wald tests) for the perceived severity model.

| Perceived susceptibility Model | | | |
|---|---|---|---|
| Predictors | Incidence rate ratios | Confidence intervals (CI) | P-value |
| household size | 1.29** | 1.28 - 1.31 | < 0.001 |
| Gender[Female]-Ref | - | - | - |
| Gender[Male] | 0.91** | 0.88 - 0.93 | <0.001 |
| Employment status [full time] - Ref | - | - | - |
| Employment status [part-time (<34 hours)] | 0.96 | 0.88 - 0.93 | 0.218 |
| Employment status [full time homemaker] | 0.80** | 0.73 -0.87 | <0.001 |
| Employment status [long term sick or disabled] | 0.71** | 0.63 - 0.81 | <0.001 |
| Employment status [Retired] | 0.70** | 0.66 - 0.75 | <0.001 |
| Employment status [Self employed] | 0.97 | 0.90 - 1.04 | 0.332 |
| Employment status [Student/pupil] | 0.89 | 0.83 - 0.96 | 0.001 |
| Employment status [Unemployed and not looking for a job] | 0.73** | 0.64 - 0.84 | <0.001 |
| Employment status [Unemployed and looking for a job] | 0.71** | 0.67 - 0.76 | <0.001 |
| High risk status [No] - Ref | | | |
| High risk status [Yes] | 0.95 | 0.92 - 0.99 | 0.006 |
| History of infection [Infected] - Ref | | | |
| History of infection [Not infected] | 1.01 | 0.94 - 1.09 | 0.785 |
| History of infection [Not tested] | 0.92 | 0.86 - 1.00 | 0.040 |
| Vaccination status [Not vaccinated] - Ref | | | |
| Vaccination status [vaccinated] | 1.22** | 1.17 - 1.27 | <0.001 |
| Perceived susceptibility [high perception] - Ref | - | - | - |
| Perceived susceptibility [low perception] | 0.91 | 0.83 - 0.99 | 0.022 |
| Perceived susceptibility [neutral perception] | 0.96 | 0.88 - 1.04 | 0.321 |
| Day of week[weekday] - Ref | | | |
| Day of week[weekend] | 0.89** | 0.86 - 0.93 | <0.001 |
| Age group [18-29] - Ref | - | - | - |
| Age group [30-39] | 0.95 | 0.90 - 1.00 | 0.045 |
| Age group [40-49] | 1.01 | 0.96 - 1.07 | 0.587 |
| Age group [50-59] | 1.03 | 0.98 - 1.09 | 0.252 |
| Age group [60-69] | 1.05 | 0.98 - 1.12 | 0.178 |
| Age group [70-120] | 1.02 | 0.93 - 1.11 | 0.745 |
| Stringency index [low] -Ref | - | - | - |
| Stringency index [Moderate] | 0.89 | 0.81 - 0.98 | 0.019 |
| Stringency index [High] | 0.82** | 0.75 - 0.90 | <0.001 |
| Stringency index [Very high] | 0.80** | 0.72 - 0.87 | <0.001 |
| Stringency index [low]* Perceived susceptibility [low perception] -Ref | - | - | - |
| Stringency index [Moderate]* Perceived susceptibility [low perception] | 0.91 | 0.80 - 1.02 | 0.109 |
| Stringency index [High]* Perceived susceptibility [low perception] | 1.03 | 0.92 - 1.16 | 0.584 |
| Stringency index [Very high]* Perceived susceptibility [low perception] | 0.95 | 0.85 - 1.06 | 0.323 |
| Stringency index [low]* Perceived susceptibility [neutral perception] -Ref | - | - | - |
| Stringency index [Moderate]* Perceived susceptibility [neutral perception] | 1.07 | 0.95 - 1.21 | 0.251 |
| Stringency index [High]* Perceived susceptibility [neutral perception] | 1.06 | 0.95 - 1.18 | 0.308 |
| Stringency index [Very high]* Perceived susceptibility [neutral perception] | 1.06 | 0.95 - 1.17 | 0.315 |

Supplementary Table 5: Results from the hierarchical generalized linear mixed effects model showing the incidence rate ratios, the associated confidence intervals and p-values for the perceived susceptibility model.

| Perceived risk to vulnerable Model | | | |
|---|---|---|---|
| Predictors | Incidence rate ratios | Confidence intervals (CI) | P-value |
| household size | 1.30** | 1.28 - 1.31 | < 0.001 |
| Gender[Female]-Ref | - | - | - |
| Gender[Male] | 0.91** | 0.88 - 0.94 | <0.001 |
| Employment status [full time] - Ref | - | - | - |
| Employment status [part-time (<34 hours)] | 0.97 | 0.91 - 1.03 | 0.267 |
| Employment status [full time homemaker] | 0.79** | 0.73 -0.86 | <0.001 |
| Employment status [long term sick or disabled] | 0.72** | 0.63 - 0.82 | <0.001 |
| Employment status [Retired] | 0.70** | 0.66 - 0.74 | <0.001 |
| Employment status [Self employed] | 0.96 | 0.90 - 1.03 | 0.300 |
| Employment status [Student/pupil] | 0.88 | 0.82 - 0.95 | 0.001 |
| Employment status [Unemployed and not looking for a job] | 0.73** | 0.63 - 0.83 | <0.001 |
| Employment status [Unemployed and looking for a job] | 0.71** | 0.67 - 0.76 | <0.001 |
| High risk status [No] - Ref | | | |
| High risk status [Yes] | 0.95 | 0.92 - 0.98 | 0.005 |
| History of infection [Infected] - Ref | | | |
| History of infection [Not infected] | 1.00 | 0.93 - 1.08 | 0.918 |
| History of infection [Not tested] | 0.92 | 0.85 - 0.99 | 0.028 |
| Vaccination status [Not vaccinated] - Ref | | | |
| Vaccination status [vaccinated] | 1.21** | 1.17 - 1.26 | <0.001 |
| Perceived risk to vulnerable [high perception] - Ref | - | - | - |
| Perceived risk to vulnerable [low perception] | 0.95 | 0.91 - 0.99 | 0.010 |
| Perceived risk to vulnerable [neutral perception] | 0.86 | 0.83 - 0.90 | < 0.001 |
| Day of week[weekday] - Ref | | | |
| Day of week[weekend] | 0.89** | 0.86 - 0.93 | <0.001 |
| Age group [18-29] - Ref | - | - | - |
| Age group [30-39] | 0.95 | 0.90 - 1.00 | 0.057 |
| Age group [40-49] | 1.02 | 0.96 - 1.07 | 0.522 |
| Age group [50-59] | 1.04 | 0.98 - 1.10 | 0.206 |
| Age group [60-69] | 1.06 | 0.99 - 1.13 | 0.114 |
| Age group [70-120] | 1.02 | 0.93 - 1.12 | 0.631 |
| Stringency index [low] -Ref | - | - | - |
| Stringency index [Moderate] | 0.88 | 0.84 - 0.93 | < 0.001 |
| Stringency index [High] | 0.84** | 0.79 - 0.90 | <0.001 |
| Stringency index [Very high] | 0.80** | 0.75 - 0.86 | <0.001 |

Supplementary Table 6: Results from the hierarchical generalized linear mixed effects model showing the incidence rate ratios, the associated confidence intervals and p-values for the perceived risk to vulnerable model.

| Perceived severity Model | | | |
|---|---|---|---|
| Predictors (Contact away from home) | Incidence rate ratios | Confidence intervals (CI) | P-value |
| household size | 1.30** | 1.28 - 1.31 | < 0.001 |
| Gender[Female]-Ref | - | - | - |
| Gender[Male] | 0.90** | 0.88 - 0.93 | <0.001 |
| Employment status [full time] - Ref | - | - | - |
| Employment status [part-time (<34 hours)] | 0.97 | 0.90 - 1.03 | 0.315 |
| Employment status [full time homemaker] | 0.79** | 0.73 -0.86 | <0.001 |
| Employment status [long term sick or disabled] | 0.71** | 0.62 - 0.81 | <0.001 |
| Employment status [Retired] | 0.69** | 0.65 - 0.74 | <0.001 |
| Employment status [Self employed] | 0.96 | 0.89 - 1.03 | 0.248 |
| Employment status [Student/pupil] | 0.87 | 0.81 - 0.94 | < 0.001 |
| Employment status [Unemployed and not looking for a job] | 0.71** | 0.62 - 0.81 | <0.001 |
| Employment status [Unemployed and looking for a job] | 0.70** | 0.66 - 0.74 | <0.001 |
| High risk status [No] - Ref | | | |
| High risk status [Yes] | 0.98 | 0.94 - 1.02 | 0.354 |
| History of infection [Infected] - Ref | | | |
| History of infection [Not infected] | 0.99 | 0.92 - 1.06 | 0.885 |
| History of infection [Not tested] | 0.91 | 0.84 - 0.98 | 0.021 |
| Vaccination status [Not vaccinated] - Ref | | | |
| Vaccination status [vaccinated] | 1.31** | 1.23 - 1.39 | <0.001 |
| Perceived severity [high perception] - Ref | - | - | - |
| Perceived severity [low perception] | 1.30** | 1.16 - 1.47 | <0.001 |
| Perceived severity [neutral perception] | 1.10 | 0.97 - 1.26 | 0.144 |
| Day of week[weekday] - Ref | | | |
| Day of week[weekend] | 0.90 | 0.87 - 0.93 | <0.001 |
| Age group [18-29] - Ref | - | - | - |
| Age group [30-39] | 1.00 | 0.91 - 1.10 | 0.982 |
| Age group [40-49] | 1.07 | 0.97 - 1.17 | 0.160 |
| Age group [50-59] | 1.06 | 0.97 - 1.16 | 0.193 |
| Age group [60-69] | 1.10 | 1.00 - 1.22 | 0.045 |
| Age group [70-120] | 1.02 | 0.91 - 1.14 | 0.778 |
| Stringency index [low] -Ref | - | - | - |
| Stringency index [Moderate] | 0.92 | 0.85 - 0.99 | 0.028 |
| Stringency index [High] | 0.87 | 0.80 - 0.95 | 0.002 |
| Stringency index [Very high] | 0.84** | 0.77 - 0.92 | <0.001 |
| Vaccination status* Perceived severity [low perception] | 0.88 | 0.80 - 0.96 | 0.003 |
| Vaccination status* Perceived severity [neutral perception] | 0.93 | 0.85 - 1.02 | 0.107 |
| Age group [18-29]* Perceived severity [low perception] -Ref | - | - | - |
| Age group [30-39]* Perceived severity [low perception] | 0.93 | 0.82 - 1.05 | 0.235 |
| Age group [40-49]* Perceived severity [low perception] | 0.91 | 0.81 - 1.03 | 0.145 |
| Age group [50-59]* Perceived severity [low perception] | 0.97 | 0.85 - 1.10 | 0.621 |
| Age group [60-69]* Perceived severity [low perception] | 0.92 | 0.80 - 1.05 | 0.205 |
| Age group [70-120]* Perceived severity [low perception] | 1.14 | 0.95 - 1.37 | 0.155 |
| Age group [18-29]* Perceived severity [neutral perception] -Ref | - | - | - |
| Age group [30-39]* Perceived severity [neutral perception] | 0.94 | 0.83 - 1.08 | 0.381 |
| Age group [40-49]* Perceived severity [neutral perception] | 0.96 | 0.85 - 1.09 | 0.557 |
| Age group [50-59]* Perceived severity [neutral perception] | 0.99 | 0.87 - 1.12 | 0.848 |
| Age group [60-69]* Perceived severity [neutral perception] | 0.92 | 0.80 - 1.05 | 0.205 |
| Age group [70-120]* Perceived severity [neutral perception] | 0.97 | 0.81 -1.15 | 0.155 |
| Stringency index [low]* Perceived severity [low perception] -Ref | - | - | - |
| Stringency index [Moderate]* Perceived severity [low perception] | 0.89 | 0.80 - 0.99 | 0.032 |
| Stringency index [High]* Perceived severity [low perception] | 0.90 | 0.81 - 1.01 | 0.083 |
| Stringency index [Very high]* Perceived severity [low perception] | 0.88 | 0.79 - 0.98 | 0.023 |
| Stringency index [low]* Perceived severity [neutral perception] -Ref | - | - | - |
| Stringency index [Moderate]* Perceived severity [neutral perception] | 1.01 | 0.90 - 1.13 | 0.927 |
| Stringency index [High]* Perceived severity [neutral perception] | 1.01 | 0.90 - 1.13 | 0.929 |
| Stringency index [Very high]* Perceived severity [neutral perception] | 0.98 | 0.87 - 1.09 | 0.701 |

Supplementary Table 7: Results from the hierarchical generalized linear mixed effects model showing the incidence rate ratios, the associated confidence intervals and p-values for the perceived severity model for the number of contacts made away from home.

| Variable | Estimate | 2.5% | 97.5% | Standard error | P-value |
|---|---|---|---|---|---|
| Intercept | 0.347 | 0.177 | 0.516 | 0.086 | <0.001 |
| Vaccination status[vaccinated]:Low | 0.074 | 0.002 | 0.147 | 0.036 | 0.042 |
| Country:Austria :Low | -.160 | -0.548 | 0.135 | 0.151 | 0.290 |
| Country:Denmark :Low | -0.433 | -0.752 | -0.154 | 0.152 | 0.002 |
| Country:Estonia :Low | 0.202 | 0.038 | 0.366 | 0.083 | 0.015 |
| Country:Finland :Low | -0.224 | -0.392 | -0.056 | 0.085 | 0.008 |
| Country:France :Low | -0.047 | -0.313 | 0.218 | 0.135 | 0.726 |
| Country:Greece :Low | 0.135 | -0.066 | 0.337 | 0.103 | 0.188 |
| Country:Italy :Low | -0.184 | -0.664 | -0.345 | 0.081 | <0.001 |
| Country:Hungary :Low | -0.504 | -0.476 | 0.108 | 0.149 | 0.217 |
| Country:Lithuania :Low | -0.502 | -0.659 | -0.344 | 0.080 | <0.001 |
| Country:Poland :Low | 0.459 | 0.155 | 0.763 | 0.155 | 0.003 |
| Country:Portugal :Low | 0.130 | -0.207 | 0.469 | 0.172 | 0.449 |
| Country:Slovakia :Low | -0.372 | -0.544 | -0.199 | 0.087 | <0.001 |
| Country:Slovenia :Low | 0.084 | -0.128 | -0.297 | 0.108 | 0.435 |
| Country:Spain :Low | 0.050 | -0.292 | 0.393 | 0.175 | 0.774 |
| Country:Switzerland :Low | -0.582 | -0.749 | -0.415 | 0.085 | <0.001 |
| Gender [male]:Low | -0.063 | | | 0.037 | 0.091 |
| Age group [30-39]:Low | 0.152 | 0.007 | 0.296 | 0.073 | 0.038 |
| Age group [40-49]:Low | 0.215 | 0.075 | 0.356 | 0.071 | 0.002 |
| Age group [50-59]:Low | 0.611 | 0.473 | 0.748 | 0.0701 | <0.001 |
| Age group [60-69]:Low | 1.071 | 0.936 | 1.206 | 0.068 | <0.001 |
| Age group [70-120]:Low | 1.551 | 1.396 | 1.707 | 0.079 | <0.001 |
| Wave[2]:Low | -0.104 | -0.164 | -0.044 | 0.0306 | <0.001 |
| Wave[3]:Low | -0.137 | -0.201 | -0.072 | 0.033 | <0.001 |
| Wave[4]:Low | -0.153 | -0.2214 | -0.085 | 0.034 | <0.001 |
| Wave[5]:Low | -0.244 | -0.316 | -0.172 | 0.0366 | <0.001 |
| Wave[6]:Low | -0.223 | -0.301 | -0.144 | 0.039 | <0.001 |
| Wave[7]:Low | -0.323 | -0.429 | -0.217 | 0.054 | <0.001 |
| Vaccination status[vaccinated]:Neutral | 0.152 | 0.060 | 0.245 | 0.047 | 0.001 |
| Country:Austria :Neutral | 0.599 | 0.292 | 0.906 | 0.156 | < 0.001 |
| Country:Denmark :Neutral | 0.212 | -0.117 | 0.542 | 0.168 | 0.206 |
| Country:Estonia :Neutral | 0.237 | 0.036 | 0.437 | 0.102 | 0.020 |
| Country:Finland :Neutral | 0.412 | 0.224 | 0.600 | 0.095 | < 0.001 |
| Country:France :Neutral | -0.091 | -0.392 | 0.210 | 0.153 | 0.553 |
| Country:Greece :Neutral | -0.118 | -0.376 | 0.139 | 0.131 | 0.368 |
| Country:Italy :Neutral | 0.060 | -0.284 | 0.406 | 0.176 | 0.729 |
| Country:Hungary :Neutral | 0.098 | -0.087 | 0.284 | 0.094 | 0.298 |
| Country:Lithuania :Neutral | -0.388 | -0.579 | -0.1989 | 0.097 | < 0.001 |
| Country:Poland :Neutral | -0.095 | -0.458 | 0.268 | 0.185 | 0.607 |
| Country:Portugal :Neutral | -0.761 | -1.265 | -0.258 | 0.257 | 0.003 |
| Country:Slovakia :Neutral | 0.400 | 0.205 | 0.595 | 0.099 | < 0.001 |
| Country:Slovenia :Neutral | 0.142 | -0.111 | 0.397 | 0.129 | 0.270 |
| Country:Spain :Neutral | 0.020 | -0.359 | 0.401 | 0.194 | 0.914 |
| Country:Switzerland :Neutral | 0.619 | 0.433 | 0.806 | 0.095 | < 0.001 |
| Gender [male]:Neutral | 0.160 | 0.079 | 0.242 | 0.041 | < 0.001 |
| Age group [30-39]:Neutral | -0.404 | -0.541 | -0.268 | 0.069 | < 0.001 |
| Age group [40-49]:Neutral | -0.526 | -0.661 | -0.391 | 0.068 | < 0.001 |
| Age group [50-59]:Neutral | -0.728 | -0.865 | -0.592 | 0.069 | < 0.001 |
| Age group [60-69]:Neutral | -0.870 | -1.010 | -0.731 | 0.070 | < 0.001 |
| Age group [70-120]:Neutral | -0.640 | -0.807 | -0.473 | 0.085 | < 0.001 |
| Wave[2]:Neutral | -0.036 | -0.112 | 0.039 | 0.038 | 0.350 |
| Wave[3]:Neutral | -0.043 | -0.124 | 0.036 | 0.041 | 0.287 |
| Wave[4]:Neutral | -0.056 | -0.142 | 0.028 | 0.043 | 0.193 |
| Wave[5]:Neutral | -0.051 | -0.141 | 0.038 | 0.046 | 0.264 |
| Wave[6]:Neutral | -0.081 | -0.179 | 0.017 | 0.050 | 0.106 |
| Wave[7]:Neutral | -0.098 | -0.234 | 0.037 | 0.069 | 0.154 |

Supplementary Table 8: Results from the generalized estimating equation model for the perceived severity for the estimated log odds of estimates and their associated 95% CI of the model variables.

| Perceived susceptibility Model | | | |
|---|---|---|---|
| Variable | Chisq | Df | Pr (> chisq) |
| Household size | 1555.942 | 1 | < 0.001 |
| Gender | 41.170 | 1 | <0.001 |
| Employment status | 255.091 | 8 | < 0.001 |
| High risk status | 7.503 | 1 | 0.015 |
| History of infection | 26.573 | 2 | <0.001 |
| Stringency index | 24.602 | 3 | <0.001 |
| Age group | 16.808 | 5 | 0.004 |
| Perceived susceptibility | 5.381 | 2 | 0.067 |
| Vaccination status | 96.101 | 1 | <0.001 |
| Day of week | 34.591 | 1 | <0.001 |
| Stringency index : Perceived susceptibility | 13.242 | 6 | 0.039 |

Supplementary Table 9: Analysis of Deviance (Type III Wald tests) for the perceived susceptibility model.

| Perceived risk to vulnerable Model | | | |
|---|---|---|---|
| Variable | Chisq | Df | Pr (> chisq) |
| Household size | 1572.45 | 1 | < 0.001 |
| Gender | 36.763 | 1 | <0.001 |
| Employment status | 260.174 | 8 | < 0.001 |
| High risk status | 7.879 | 1 | 0.005 |
| History of infection | 26.322 | 2 | <0.001 |
| Stringency index | 48.940 | 3 | <0.001 |
| Age group | 17.560 | 5 | < 0.001 |
| Perceived risk to vulnerable | 49.083 | 2 | < 0.001 |
| Vaccination status | 91.285 | 1 | <0.001 |
| Day of week | 34.640 | 1 | <0.001 |

Supplementary Table 10: Analysis of Deviance (Type III Wald tests) for the perceived risk to vulnerable model.

| Country | Cronbach's alpha | 95% Confidence intervals |
|---|---|---|
| Austria | 0.61 | 0.59 - 0.62 |
| Croatia | 0.67 | 0.65 - 0.68 |
| Denmark | 0.33 | 0.30 - 0.36 |
| Estonia | 0.67 | 0.66 - 0.69 |
| Finland | 0.65 | 0.63 - 0.66 |
| France | 0.70 | 0.68 - 0.71 |
| Greece | 0.74 | 0.73 - 0.75 |
| Hungary | 0.67 | 0.66 - 0.69 |
| Italy | 0.64 | 0.63 - 0.66 |
| Lithuania | 0.76 | 0.75 - 0.77 |
| Poland | 0.75 | 0.74 - 0.76 |
| Portugal | 0.41 | 0.38 - 0.43 |
| Slovakia | 0.70 | 0.69 - 0.71 |
| Slovenia | 0.65 | 0.63 - 0.66 |
| Spain | 0.56 | 0.54 - 0.58 |
| Switzerland | 0.66 | 0.65 - 0.68 |

Supplementary Table 11: Cronbach's alpha for each individual country.
